# Intersectional inequalities in youth mental health measurement and outcomes: population-representative evidence from the UK Millennium Cohort Study

**DOI:** 10.64898/2026.10.19.26365893

**Authors:** Darío Moreno-Agostino, Thomas Steare, Clare R. Evans, Emla Fitzsimons, Ann Phoenix, George B. Ploubidis, Usayd Abid, Yusra Nawaz, Hemlata Pant, Helen L. Fisher, Praveetha Patalay

## Abstract

**Background:** Population-representative evidence on social inequalities in youth mental health from an intersectional perspective is limited. We investigated inequalities across multiple mental health outcomes among young adults and examined whether measures operated equivalently across social and intersectional groups.

**Methods:** Cross-sectional analyses were conducted with 9,675 participants aged 23 years from the nationally representative UK Millennium Cohort Study (born 2000-2002). Self-reported outcomes included psychological distress (Kessler-6), probable serious psychological distress/‘caseness’ (Kessler-6 score ≥13), psychological wellbeing (Short Warwick-Edinburgh Mental Well-Being Scale; SWEMWBS), past-year self-harm, and lifetime suicide attempts. Measurement invariance analyses assessed comparability of the Kessler-6 and SWEMWBS across groups. Intersectional MAIHDA examined inequalities by sex assigned at birth, ethnic identity/background, sexual identity, and childhood poverty. Young people with lived experience of mental ill-health and/or social discrimination contributed to study design, interpretation, and reporting.

**Findings:** Large inequalities were observed across all outcomes, with the largest disparities by sexual identity, followed by sex assigned at birth, childhood poverty history, and ethnic identity/background. Across intersectional strata, predicted psychological distress ranged from-0.27 to 1.32 SDs, and psychological wellbeing from-0.54 to 0.11 SDs. Predicted prevalence varied 6.99-fold (9.81% to 68.55%) for psychological distress ‘caseness’, 6.84-fold (8.60% to 58.79%) for self-harm, and 22.10-fold (1.96% to 43.32%) for suicide attempts. Young people at the intersection of sexual minority status, female birth sex, childhood poverty, and some ethnic minority identities/backgrounds experienced particularly poor outcomes. Most inequalities showed cumulative patterns consistent with additive expectations, although some intersectional groups experienced better or worse outcomes than expected. Accounting for measurement differences revealed inequalities that were otherwise undetected.

**Interpretation:** Findings highlight the large extent of mental health inequalities when multiple dimensions of social (dis)advantage are considered, and the need to address both the social conditions generating these inequalities and the methodological assumptions shaping how they are measured and understood.

## Introduction

Mental ill-health is among the top global contributors to disability, morbidity, and mortality in young people.^1–3^ In England, the most recent population estimates (2023/2024) suggest that around 25.8% of people aged 16-24 years have a probable mental disorder.^4^ Youth mental health is socially patterned: young women and sexual minority young people report substantially poorer mental health outcomes and, in the case of sexual and gender minorities, much higher self-harm and suicide rates than their heterosexual peers.^5–8^ Young people from disadvantaged socioeconomic backgrounds also experience worse mental health outcomes,^9–11^ with mental health inequalities by childhood socioeconomic position (SEP) still evident more than 50 years later.^12^ Evidence on ethnic inequalities is more complex, with some UK studies reporting higher levels of mental health symptoms among youth from White and Mixed ethnic backgrounds.^6,13^

These inequalities arise through interlocking systems of oppression and disadvantage, including sexism, racism, heterosexism, classism, and socioeconomic inequality, which shape exposure to discrimination, access to resources, and opportunities for good mental health. Rooted in Black feminist scholarship and activism, an intersectional framework highlights that these systems are not independent but mutually constitutive.^14–16^ Quantitatively, intersectional inequalities may reflect cumulative patterns across social positions, with some groups departing from additive expectations. Without an explicit intersectional approach, such inequalities will remain invisible.^17^

Despite growing recognition of intersectional complexity in mental health research,^18^ population-representative evidence on intersectional inequalities in youth mental health in the UK remains limited. Existing studies also typically assume measurement comparability across social groups, despite the possibility that mental health measures may not function equivalently across groups, potentially biasing estimates of mental health inequalities.^19^ Furthermore, most studies focus on single outcomes, even though different indicators capture distinct aspects of mental health and may be socially patterned in different ways.^7,20^

In this preregistered study, shaped by the involvement of young people with lived experience of mental ill-health and/or social discrimination, we addressed two objectives. First, we examined whether commonly used measures of mental ill-health and wellbeing were comparable across multiple social identities and positions, and their intersections. Second, we investigated inequalities in multiple mental health outcomes across intersectional strata and assessed whether these inequalities departed from additive expectations associated with the social identities and positions considered. We expected that some indicators would show differential measurement properties across groups, and that most inequalities would reflect cumulative patterns consistent with additive expectations, while some would reflect intersection-specific inequalities (interactions).

## Methods

### Study design and participants

In this cross-sectional study, we used data from the Millennium Cohort Study (MCS), a population-based UK birth cohort of 19,517 individuals born between 2000 and 2002.^21,22^ Children from disadvantaged areas, minoritised ethnic backgrounds, and the smaller UK nations were oversampled. Participants have been followed from age 9 months to 23 years through eight survey sweeps. In this study, we included 9,675 non-proxy respondents who participated in the age 23 sweep.^23^

The age 23 MCS sweep used a sequential mixed-mode design: online survey, followed by face-to-face interviews for non-respondents, and a shortened online-only mop-up survey for remaining non-respondents (making up around 4.7% of the productive interviews). Participants who had withdrawn their consent for use of their data and triplets (whose data are not available in the End User License dataset) were excluded. Ethical approval for MCS data collection was obtained from the relevant NHS Research Ethics Committees, and this study received approval from the UCL IOE Research Ethics Committee (REC2074). This study is reported in accordance with STROBE guidelines (**Appendix 1, pp3-5)**.

### Measures

#### Outcomes

We examined five mental health outcomes based on information self-reported at age 23 years: psychological distress measured with the Kessler-6,^24^ ‘caseness’ (i.e., probable serious psychological distress, Kessler-6 score ≥13),^25^ psychological wellbeing measured with the Short Warwick-Edinburgh Mental Well-Being Scale (SWEMWBS),^26^ self-harm in the past 12 months,^27^ and lifetime suicide attempts. For participants reporting any suicide attempts at age 17 but not age 23, the age-17 response was retained. Additional details on the outcome measures are provided in **Appendix 1 (p6)**.

#### Social identities and positions

Consistent with an inter-categorical approach to intersectionality,^28^ we used data on four social identities and positions as proxies for the systems of oppression under study (sexism, racism, heterosexism, and classism and the material effects of early-life economic deprivation). These were: sex assigned at birth (1=female, 2=male), ethnic identity (1=White, 2=Mixed, 3=Indian, 4=Pakistani/Bangladeshi, 5=Black, 6=Other), sexual identity (0=heterosexual, 1=sexual minority), and childhood poverty (household income: 0=never, 1=ever lived in a household with income below 60% of the national OECD-equivalised median during early childhood).^9,10^ These variables were combined to define up to 48 intersectional strata representing different combinations of social identities and positions. Additional details on the variable construction and category definitions are provided in **Appendix 1 (pp7-8)**.

## Statistical analysis

We first used the alignment method,^29^ an optimisation-based measurement approach, to test whether the Kessler-6 and SWEMWBS measured the same underlying constructs across groups. Measures were considered sufficiently comparable if no more than 25% of the measurement parameters were non-invariant (with α=0.001).^30^ Where more extensive non-invariance was identified, we used multiple-group confirmatory factor analysis (MG-CFA) and partial measurement invariance models to identify and account for non-invariant item parameters.^30^ Factor scores (mean=0, SD=1) derived from the partially invariant models were used in the main analyses to account for any identified measurement differences across groups, and analyses with the observed sum scores were conducted for comparison purposes (see Sensitivity analyses). See **Appendix 1 (pp9-11)** for further details.

Next, we used I-MAIHDA (Intersectional Multilevel Analysis of Individual Heterogeneity and Discriminatory Accuracy) models,^31,32^ with individuals nested within the intersectional strata defined by combinations of social identities and positions. Linear multilevel models were fitted for continuous outcomes and logistic multilevel models for binary outcomes. We first estimated intercepts-only (null) models to quantify between-stratum inequalities using the variance partition coefficient (VPC_null_). We then fitted models including the main/additive effects of the variables defining the strata and calculated the proportional change in between-stratum variance (PCV). Remaining between-stratum variance after accounting for main/additive effects was interpreted as evidence of departures from additive expectations (non-additive/interaction effects), with smaller PCVs indicating greater remaining between-stratum variation. To characterise these departures, we examined stratum-specific empirical Bayes residuals from the main-effects models. Predicted means (continuous outcomes) and probabilities (binary outcomes) were derived for each intersectional stratum to describe patterns of inequality. See **Appendix 1 (p12)** for further details.

To restore representativeness to the target population (individuals born in the UK between 2000 and 2002 and alive at age 23 years), all analyses were weighted using combined survey and non-response weights that account for unequal selection probabilities and differential participation in the age 23 sweep.^23^ Analyses were restricted to participants with complete data on the variables included in each model. Following Office for National Statistics disclosure control guidance, results for strata with fewer than 10 observations were suppressed.

### Sensitivity analyses

First, we replicated the analyses using the Kessler-6 and SWEMWBS observed sum scores to assess the potential impact on the results of ignoring the identified measurement differences across groups. Second, because the MCS mop-up survey included the Kessler-6 but not the other mental health measures, sensitivity analyses for psychological distress and ‘caseness’ excluded mop-up respondents to have comparable sample across outcomes. Third, we conducted quantitative bias analyses (QBA) to assess the potential impact of selection and mode effects between online and face-to-face respondents (additional details provided in **Appendix 1, p13**).^33^ Finally, to assess the robustness of findings to outcome definition, self-harm analyses were conducted using the number of self-harm behaviours (count) rather than any self-harm (binary), using negative binomial and Poisson I-MAIHDA models (**Appendix 1, p12**).

Data management and I-MAIHDA models were conducted in Stata/MP version 19.5.^34^ Measurement invariance analyses were conducted in Mplus version 9.1.^35^ Data management and analysis code is publicly available in the GitHub (https://github.com/dmorenoagostino/intersectymh-mcs8).

### Lived experience involvement

A Young People Advisory Group (YPAG) comprised of 13 young people from diverse intersectional positions with lived experience of mental ill-health and/or social discrimination shaped the research question, study design, and protocol, and helped interpret the results and prepare the manuscript. See **Appendix 1 (p14)** for further details.

### Role of the funding source

The study funders had no role in study design; data collection, analysis, or interpretation; writing of the report; or the decision to submit the paper for publication.

The study was preregistered in the Open Science Framework (OSF, https://doi.org/10.17605/OSF.IO/U89GR). Deviations from the preregistered protocol are described in **Appendix 1 (pp15-16)**.

## Results

### Participant characteristics

Analyses included 9,675 participants aged 23 years, of whom just over half were female, around four in five identified as White, one in six belonged to a sexual minority group, and two in five had experienced childhood poverty (**Table 1**). Most participants completed the survey online. Weighted mean scores were 8.4 for the Kessler-6 and 23.3 for the SWEMWBS. The prevalence of ‘caseness’/probable psychological distress, past-year self-harm, and lifetime suicide attempt was 21.3%, 20.3%, and 13.1%, respectively. Missingness was low across outcomes (3.0%-3.5%; **Table 1**). Of the 48 intersectional strata, 13 (27.1%) comprised at least n=100 participants, 30 (62.5%) at least n=20, and 16 (33.3%) had less than 10 observations, with one of those having none. All intersectional strata with less than 10 observations included sexual minority youth from ethnically minoritised groups. **Table 1** presents the weighted population distribution of social identities and positions and corresponding weighted outcome estimates. The visualisation of these descriptive statistics along additional details at the more granular ethnic and sexual identity categories before recoding are available in **Appendix 1 (pp17-22)**.

**Table 1.**
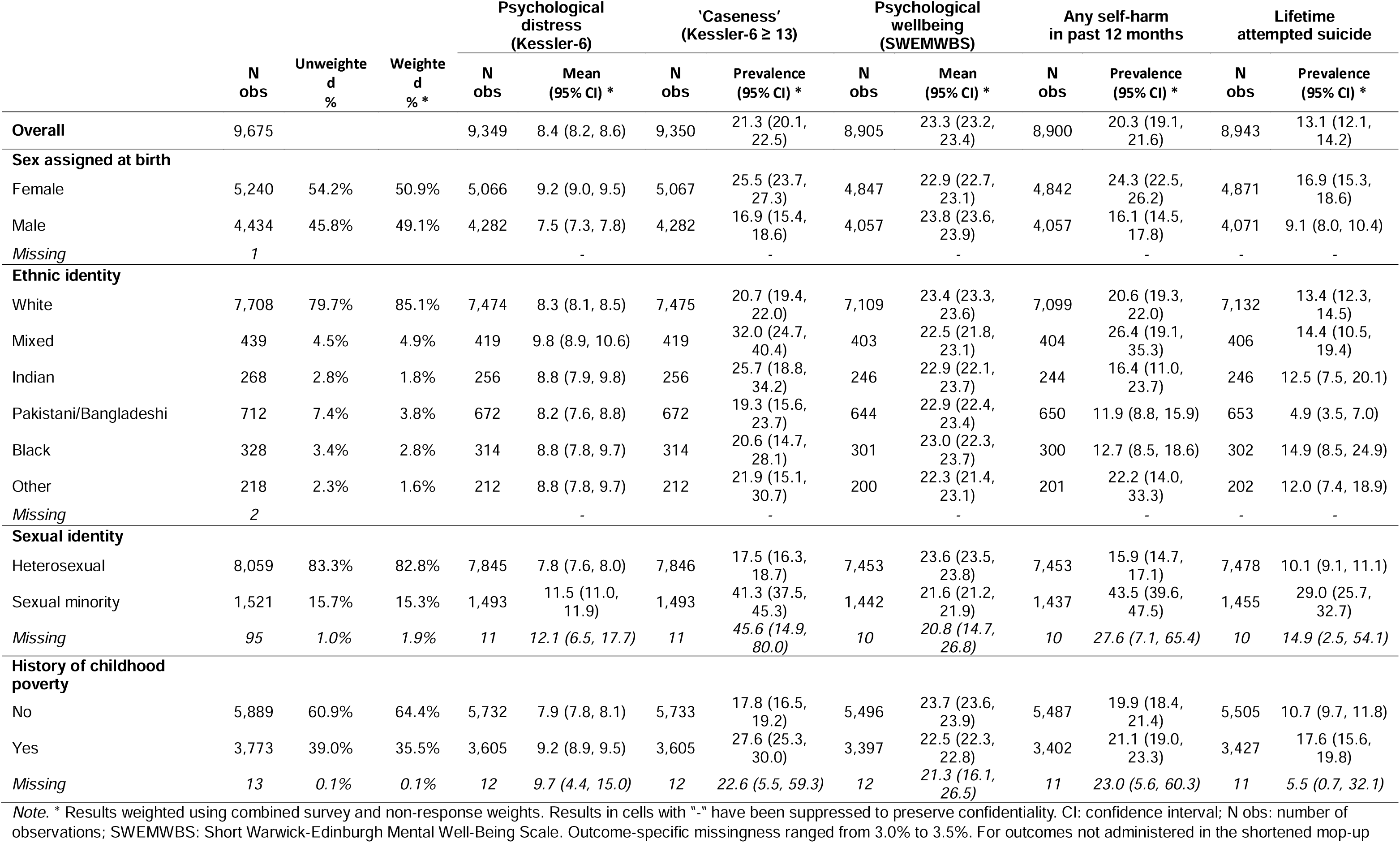
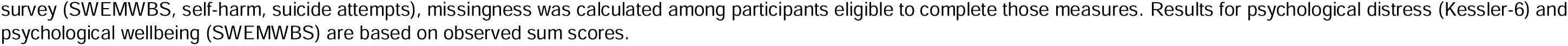
Distribution of social identities and positions and outcome estimates.

### Measurement invariance

Initial alignment analyses suggested measurement non-invariance by sexual identity for the Kessler-6 (58.3% of parameters flagged as non-invariant) and by birth sex for the SWEMWBS (28.6%; **Appendix 1, pp23-27**). Subsequent MG-CFA identified no comparability issues for the Kessler-6 across sexual identity alone; however, analyses further incorporating birth sex, childhood poverty, and their intersections consistently indicated partial measurement non-invariance. At the most granular level examined (birth sex * sexual identity * childhood poverty), non-invariance in the Kessler-6 was concentrated in three items (“nervous”, “restless or fidgety”, “everything was an effort”), whereas non-invariance in the SWEMWBS was limited to three items (“feeling optimistic about the future”, “feeling close to other people”, and “able to make up my own mind”) among heterosexual females without a history of childhood poverty. Partially invariant factor scores were subsequently derived from the final measurement models for use in the main analyses.

### Social inequalities in mental health outcomes

We found substantial additive inequalities across all outcomes (**Figure 1**). Sexual minority status was associated with the largest inequalities, with consistently poorer mental health and wellbeing. Compared with heterosexual participants, sexual minority youth reported higher psychological distress (mean difference in SD units 0.66 [95% CI: 0.53, 0.79]), lower wellbeing (mean difference in SD units-0.27 [95% CI:-0.34,-0.20]), and more than threefold higher odds of self-harm (OR 4.21 [95% CI: 3.11, 5.69) and lifetime suicide attempt (OR 3.39 [95% CI: 3.09, 3.71]).

**Figure 1.**
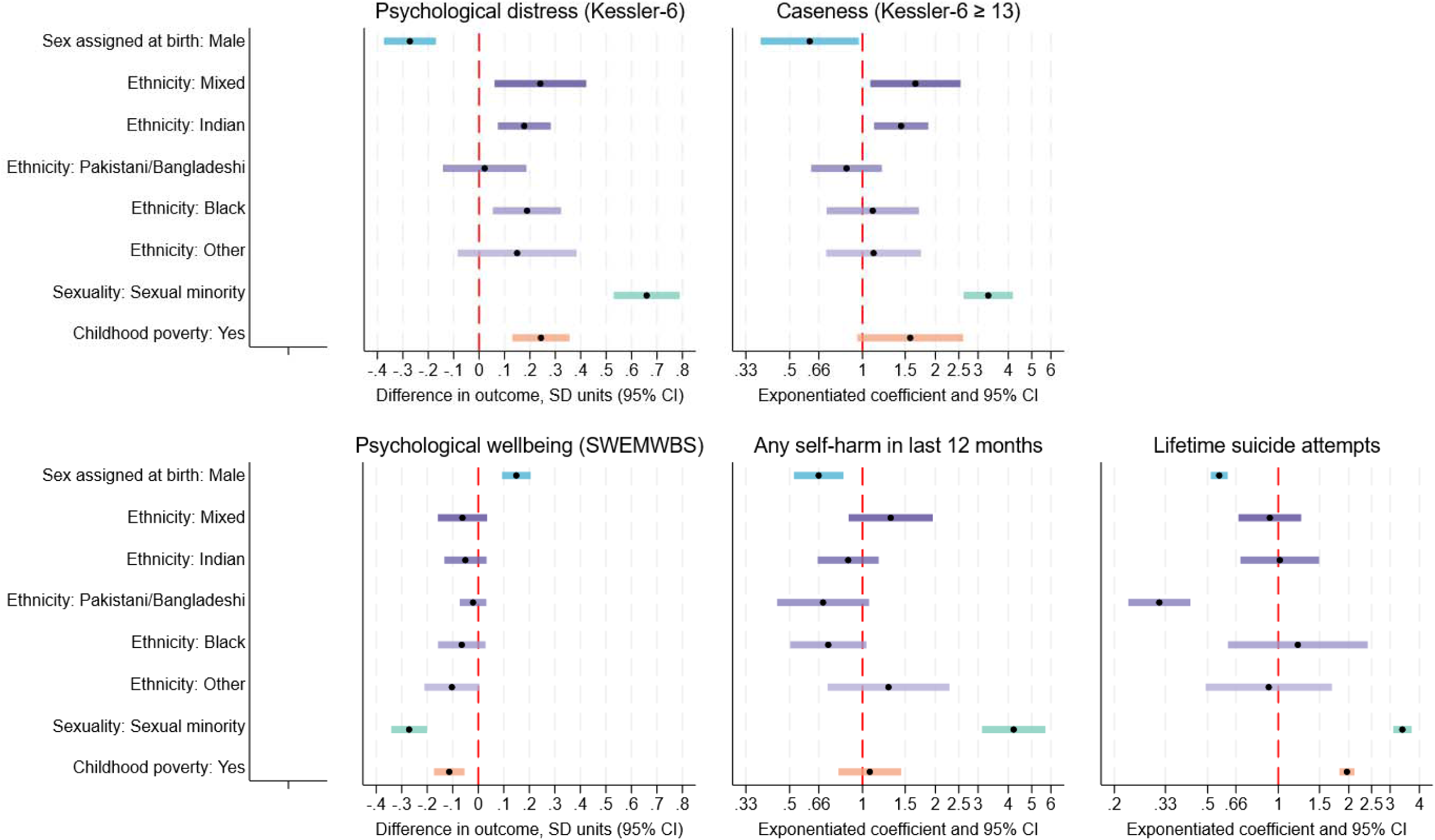
Coefficient plot of additive effects across outcomes, weighted results. *Note*. CI: confidence interval; SD: standard deviation; SWEMWBS: Short Warwick Edinburgh Mental Well-Being Scale. The red dashed line represents the absence of effect, located at 0 for continuous outcomes and 1 for binary outcomes. Black dots represent the point estimate for each effect, and accompanyi coloured line represents the 95% confidence interval (CI). Additive effects have been calculated using Sex assigned at birth: Female, Ethnicity: White, Sexu Heterosexual, and Childhood poverty: No as reference categories. Results for psychological distress (Kessler-6) and psychological wellbeing (SWEMWBS) are based on partially invariant factor scores. Results weighted using combined survey and non-response weights.

Females also reported worse outcomes than males across all outcomes, although inequalities were generally smaller than those observed by sexual identity. Childhood poverty was associated with poorer mental health and wellbeing, with particularly large inequalities in lifetime suicide attempts (OR 1.96 [95% CI: 1.82, 2.11]). Ethnic inequalities were more heterogeneous. Compared with White participants, those from Mixed, Indian, and Black ethnic identities/backgrounds reported higher psychological distress. Participants from Pakistani or Bangladeshi ethnic backgrounds had substantially lower odds of lifetime suicide attempts (OR 0.31 [95% CI: 0.23, 0.42]).

In intercepts-only (null) I-MAIHDA models, VPC_null_ estimates ranged from 10.3% for psychological wellbeing to 20.4% for self-harm, indicating that the degree of clustering within intersectional strata varied across outcomes (**Table 2**). PCV estimates ranged from 87.4% for wellbeing to 100% for lifetime suicide attempts, indicating that most between-strata variation was accounted for by additive differences associated with sex assigned at birth, ethnic identity/background, sexual identity, and childhood poverty. Lifetime suicide attempts showed high initial clustering but no remaining between-strata variation after adjustment for these additive effects, whereas psychological wellbeing showed the lowest initial clustering and the largest remaining proportion of unexplained between-strata variation.

**Table 2.** Variance partition coefficient (VPCs) and proportional change in variance (PCV) estimates from main I-MAIHDA models, weighted results.

| Outcome | N | VPC<sub>null</sub> (95% CI) | PCV |
| --- | --- | --- | --- |
| Kessler-6, factor scores | 9326 | 16.7 (9.1, 28.5) | 91.9 |
| 'Caseness' (Kessler-6 $\geq 13$) | 9327 | 15.1 (7.7, 27.5) | 95.3 |
| SWEMWBS, factor scores | 8883 | 10.3 (5.7, 18.1) | 87.4 |
| Self-harm (any) | 8879 | 20.4 (12.9, 30.8) | 96.3 |
| Lifetime suicide attempts | 8922 | 20.0 (13.4, 28.9) | 100 |

*Note.* CI: confidence interval; I-MAIHDA: intersectional multilevel analysis of individual heterogeneity and discriminatory accuracy; SWEMWBS: Short Warwick-Edinburgh Mental Well-Being Scale; VPC: variance partition coefficient; PCV: proportional change in variance. VPC<sub>null</sub> was estimated from intercepts-only models and it represents the proportion of the total variance in each mental health outcome attributable to differences between, rather than within, intersectional strata, here expressed as a percentage. PCV is the proportional reduction in between-strata variance between the intercepts-only (null) and main-effects models, expressed as a percentage. Results weighted using combined survey and non-response weights.

Examination of the remaining between-strata variation identified several intersectional strata with mental health outcomes better or worse than cumulative (additive) expectations (**Figure 2**). White females with no history of childhood poverty had better-than-expected psychological distress and wellbeing, with slightly larger departures among sexual minority than heterosexual youth (distress:-0.18 [95% CI:-0.24,-0.11] vs-0.07 [-0.11,-0.04]; wellbeing: 0.09 [0.06, 0.13] vs 0.04 [0.02, 0.06]). Heterosexual males from a Mixed ethnic identity/background with a history of childhood poverty also had better-than-expected psychological distress (-0.17 [-0.32,-0.03]). Conversely, White heterosexual males with no history of childhood poverty had slightly worse-than-expected psychological distress (0.06 [0.02, 0.09]). Using partially invariant factor scores allowed identifying worse-than-expected outcome levels that could not be detected using the observed sum scores (**Appendix 1, p29**) among White sexual minority youth with a history of childhood poverty, both in males (distress: 0.12 [0.01, 0.23], wellbeing:-0.07 [-0.13,-0.01]) and females (wellbeing:-0.06 [-0.12,-0.01]).

**Figure 2.**
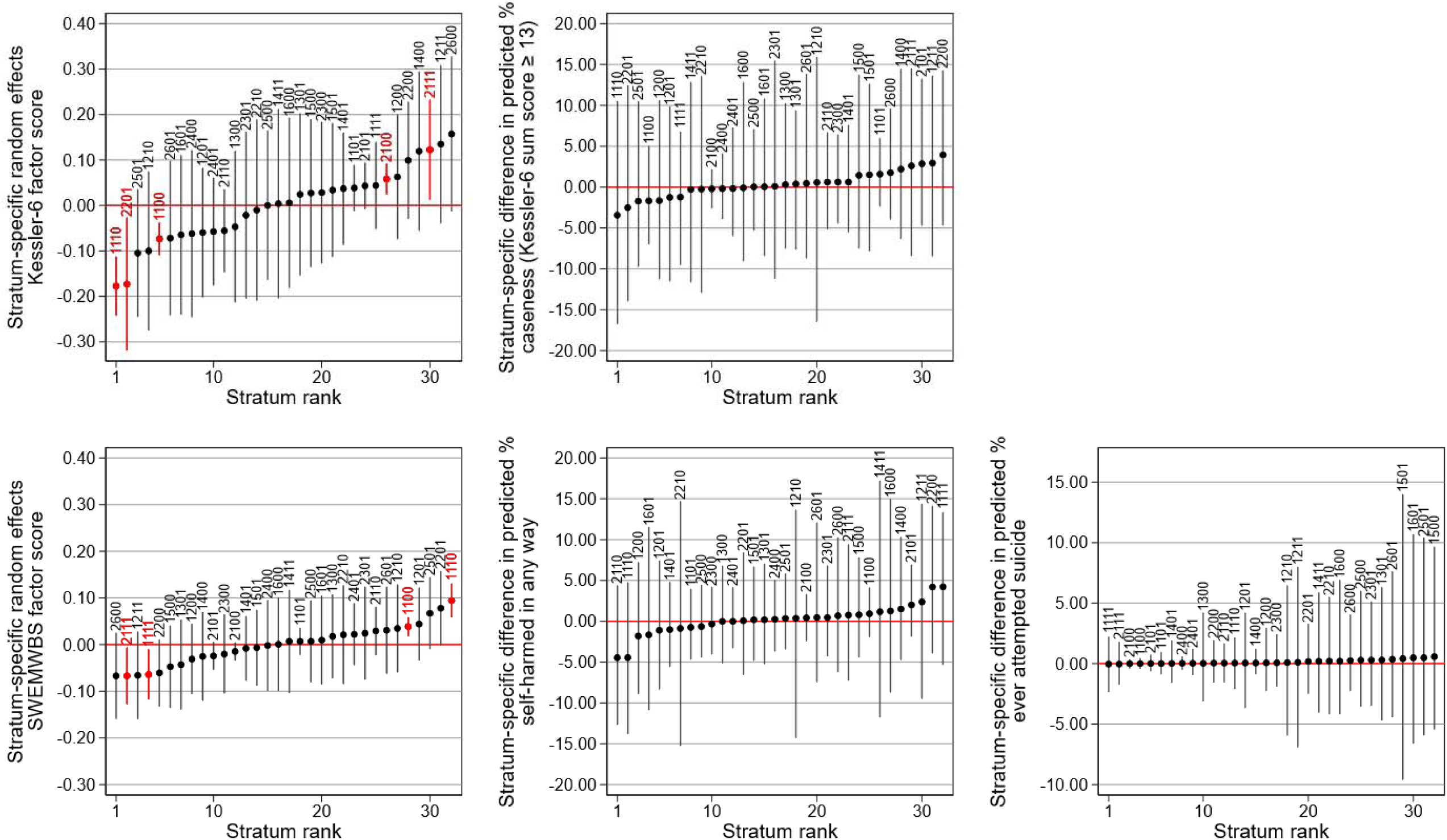
Caterpillar plots of random (non-additive) effects across intersectional strata, weighted results. *Note.* SWEMWBS: Short Warwick Edinburgh Mental Well-Being Scale. Dots represent random (non-additive) effects (expressed as the difference in mean prevalence estimates compared to the expectation based on additive effects alone), and accompanying lines represent the 95% confidence intervals. Rand effects significantly different than zero (not overlapping with zero) are highlighted in red. Intersectional strata are coded as follows: first digit: sex assigned a (1 female, 2 male); second digit: ethnic identity/background (1 White, 2 Mixed, 3 Indian, 4 Pakistani/Bangladeshi, 5 Black, 6 Other); third digit: sexual identit Heterosexual, 1 Sexual minority); fourth digit: history of childhood poverty (0 No, 1 Yes). Results from strata with less than 10 observations are not shown to preserve confidentiality. Results for psychological distress (Kessler-6) and psychological wellbeing (SWEMWBS) are based on partially invariant factor scores. Results weighted using combined survey and non-response weights.

Taken together, cumulative inequalities across sex assigned at birth, ethnic identity/background, sexual identity, and childhood poverty, alongside smaller departures from additive expectations in some strata, produced substantial variation in predicted mental health levels across intersectional positions (**Figure 3**). Among non-suppressed strata, predicted psychological distress ranged from-0.27 to 1.32 SDs (difference 1.59 SDs), whereas predicted psychological wellbeing ranged from-0.54 to 0.11 SDs (difference 0.65 SDs). Predicted prevalence ranged from 9.81% to 68.55% for ‘caseness’/probable psychological distress (difference 58.74 percentage points; 6.99-fold), 8.60% to 58.79% for self-harm (difference 50.19 percentage points; 6.84-fold), and 1.96% to 43.32% for lifetime suicide attempts (difference 41.36 percentage points; 22.10-fold).

**Figure 3.**
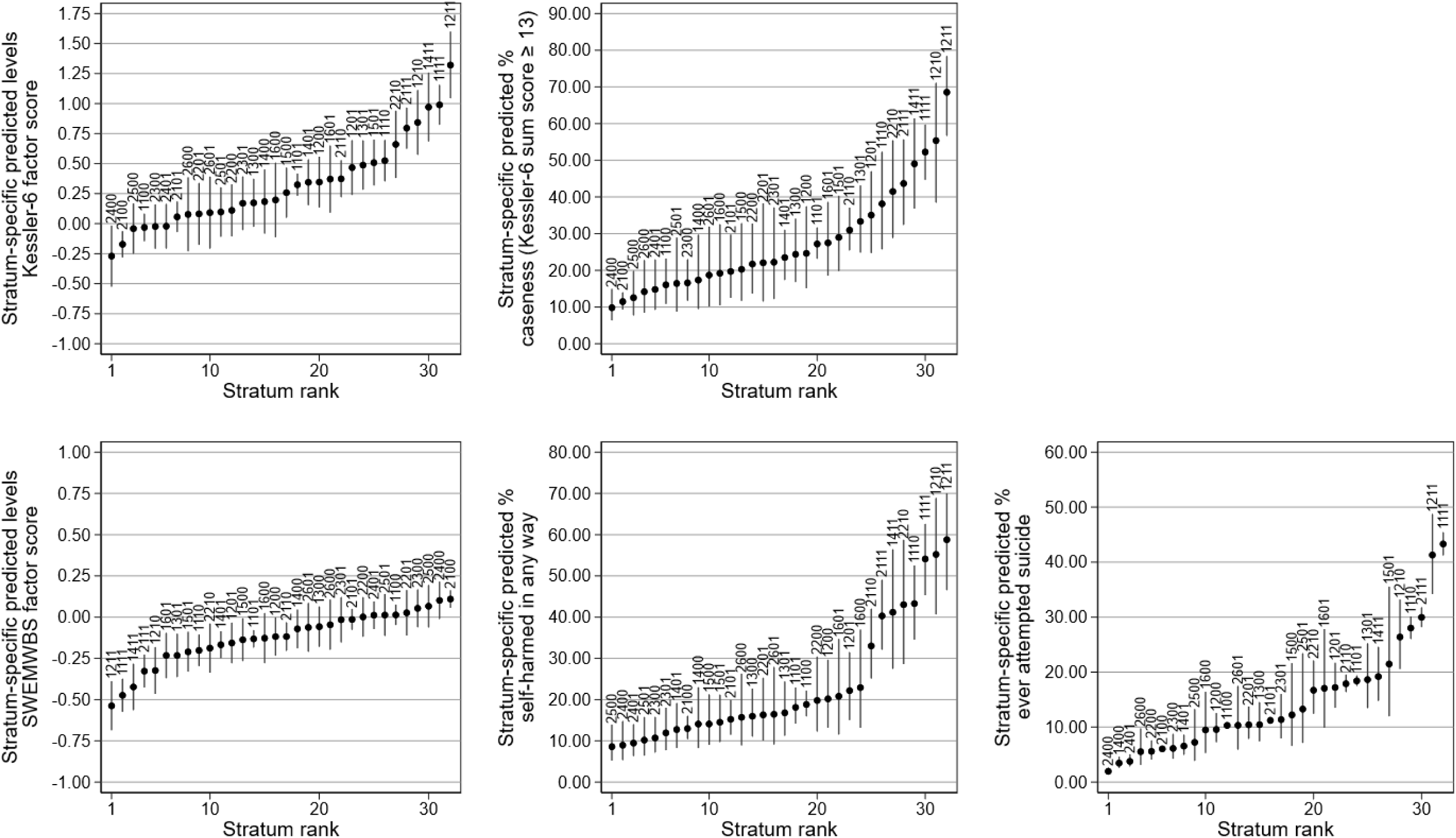
Caterpillar plots of predicted outcome levels across intersectional strata, weighted results. *Note.* Dots represent predicted outcome levels (means for Kessler-6 and Short Warwick Edinburgh Mental Well-Being Scale (SWEMWBS), prevalence esti for ‘caseness’, self-harm, and lifetime suicide attempts), and accompanying lines represent the 95% confidence intervals. Intersectional strata are coded as follows: first digit: sex assigned at birth (1 female, 2 male); second digit: ethnic identity/background (1 White, 2 Mixed, 3 Indian, 4 Pakistani/Bangladeshi, 5 B 6 Other); third digit: sexual identity (0 Heterosexual, 1 Sexual minority); fourth digit: history of childhood poverty (0 No, 1 Yes). Results from strata with less t 10 observations are not shown to preserve confidentiality. Results for psychological distress (Kessler-6) and psychological wellbeing (SWEMWBS) are based on partially invariant factor scores. Results weighted using combined survey and non-response weights.

Across outcomes, the poorest predicted mental health outcomes were generally observed among sexual minority females with a history of childhood poverty, particularly those from Mixed ethnic backgrounds. Conversely, the most favourable predicted outcomes were predominantly observed among heterosexual youth without a history of childhood poverty, particularly males from White or Pakistani or Bangladeshi backgrounds.

### Sensitivity analyses

Beyond the differences between analyses using partially invariant factor scores and observed sum scores already described, most findings were largely unchanged across sensitivity analyses (**Appendix 1, pp28-35**), with a small number of exceptions. Worse-than-expected distress among White heterosexual males with a history of childhood poverty was significant only when observed sum scores were used. Most remaining differences emerged across QBA scenarios. Under the most extreme QBA scenario, non-additive effects for wellbeing were completely attenuated. Worse-than-expected distress and wellbeing among White sexual minority males with a history of childhood poverty were not significant in several QBA scenarios. Likewise, better-than-expected distress among heterosexual males from a Mixed ethnic identity/background with a history of childhood poverty was not significant in the most extreme QBA scenario, whereas better-than-expected wellbeing in this stratum emerged in the least extreme QBA scenario. Better-than-expected distress among White sexual minority females with a history of childhood poverty was no longer significant in the two most extreme QBA scenarios.

Analyses using the number of self-harm behaviours rather than any self-harm showed a broadly similar pattern of inequalities. Although Poisson models suggested additional non-additive effects, these were not replicated in negative binomial models and were therefore not considered robust. Model-wide and stratum-specific estimates and rankings for all estimated models are provided in **Appendix 2**.

## Discussion

In this nationally representative study of young adults born in the UK in the early 2000s, we found substantial social inequalities across multiple mental health outcomes. For example, the predicted prevalence of self-harm and lifetime suicide attempts varied more than six-fold and twenty-fold, respectively, across intersectional strata. These inequalities showed strong cumulative patterns across dimensions of social (dis)advantage largely consistent with additive expectations, with sexual minority status consistently associated with the largest disparities across all outcomes. Young people at the intersection of sexual minority status, female birth sex, childhood poverty, and certain ethnic identities/backgrounds (most frequently those with a Mixed ethnic identity/background) were at particularly high risk of poor mental health outcomes, largely reflecting the cumulative patterning of these inequalities. Nevertheless, some intersectional strata showed mental health outcomes that departed from these cumulative (additive) expectations, and some of these departures were only detectable after accounting for systematic differences in how psychological distress and wellbeing were measured across intersectional strata.

We found measurement differences in psychological distress and wellbeing across intersectional strata. Because these differences were generally modest and most indicators remained invariant across strata, we were able to derive factor scores that incorporated both the commonalities and the identified differences in measurement, providing a more comparable characterisation of mental health across groups. Our findings highlight a largely overlooked dimension of intersectional complexity in quantitative research.^36^ The prevalence and consequences of measurement differences in other populations, contexts, and measures remain largely unknown. Just as intersectional inequalities in the outcomes can remain hidden unless explicitly examined,^17^ systematic differences in the measurement of those outcomes may also obscure, exaggerate, or otherwise distort observed inequalities. In the context of youth mental health research, such differences are likely as the development and validation of mental health measures rarely involve meaningful engagement with the diverse communities and intersectional groups to whom they are subsequently applied.^37^

We found consistent and robust evidence that sexual minority youth experienced the largest disadvantages in mental health, reporting substantially higher distress, lower wellbeing, and at least threefold higher odds of self-harm and lifetime suicide attempts than their heterosexual counterparts. This is consistent with initial descriptive findings from the same cohort at age 23,^38^ as well as previous evidence from earlier in the life courses and other populations.^6,20,39^ Our findings suggest that heterosexism continues to shape the lives and mental health of sexual minority young adults in the UK despite this being a life stage when greater autonomy may facilitate experiences of affirmation, belonging, and queer joy.^40^ Although with gaps smaller than by sexual identity, young adults assigned female at birth also experienced substantial disadvantages across all outcomes, pointing at the persisting impacts of sexism and misogyny on young people from this generation.^6,20,41^

Unlike previous results from the same birth cohort at age 17,^20^ our results show that White youth in their early twenties no longer report the worst mental health outcomes. Although our study is not longitudinal, this contrast with earlier findings suggests that the configuration of intersectional inequalities may change across developmental periods and social contexts. These findings also align with documented inequalities experienced by ethnic minority adults in the UK in access to mental health care, experiences through care, and outcomes due to (institutional) racism,^42,43^ highlighting the need to better understand how intersectional inequalities emerge and change over time.

Having grown up in a household below the poverty line was related with twice the odds of having ever attempted a suicide, with more modest inequalities in other outcomes. This highlights the importance of using multiple outcomes when investigating population mental health and also points at the lasting impacts of early life economic deprivation and its role as a fundamental cause of inequalities across the life course.^44^ Together, these inequalities combined to produce a highly unequal picture of mental health in youth, including a more than 40 percentage-point gap in the predicted prevalence of lifetime suicide attempts between the highest-and lowest-risk groups.

Among White youth without a history of childhood poverty, females had better-than-expected psychological distress and wellbeing outcomes, particularly among sexual minority youth, whereas heterosexual males had worse-than-expected psychological distress. Although our study was not designed to identify the mechanisms underlying these departures from additive expectations, our interpretation, informed by discussions with the YPAG, considered potential differences in social support, help-seeking, community connectedness, and access to affirming environments during the transition to adulthood.

The somewhat buffered outcomes observed among some sexual minority females, and in heterosexual males with a Mixed ethnic identity/background and history of childhood poverty may also reflect resilience developed through navigating marginalisation earlier in life and, in the case of sexual minority youth, experiences of belonging or queer joy that are not captured in our analyses.^40^ Conversely, worse-than-expected outcomes among some White males may reflect the influence of masculine norms discouraging emotional expression and support seeking.^45^ Additional worse-than-expected patterns observed among White sexual minority youth with a history of childhood poverty after accounting for measurement differences may point to forms of disadvantage not captured in our study, including potential additional barriers to disclosure.^46^

To our knowledge, this is the first study to examine social and intersectional inequalities in youth mental health while explicitly acknowledging intersectional complexity in both the outcomes and the way many of them are measured, and to incorporate youth involvement throughout the research process. Our findings provide an updated, inclusive, and more accurate characterisation of social inequalities in multiple mental health outcomes at the intersection of sexism, racism, heterosexism, and classism and early-life socioeconomic deprivation.

However, several limitations must be considered. Due to data constraints, we grouped together some ethnic and sexual identity categories with distinct social, cultural, and historical experiences of marginalisation, reducing the granularity of our analyses. Although descriptive statistics for more detailed categories are presented, this level of detail could not be maintained in the intersectional analyses. Even after aggregation, several intersectional strata comprised fewer than 10 individuals so estimates could not be reported to preserve confidentiality. All these strata included combinations of ethnic and sexual minority identities/backgrounds and were often among those with the poorest predicted mental health outcomes (**Appendix 2**). Limited sample sizes may also have reduced our ability to detect non-additive effects in some strata. Future studies with larger samples, potentially through pooled datasets, may help address these challenges.

We used social identities and positions as proxies for the systems of oppression whose impacts on mental health inequalities we aimed to investigate.^28^ As such, our findings identify where inequalities occur rather than the mechanisms that produce them. Additive effects were conditional on the other social identities/positions included in the models and should therefore not be interpreted as total effects, particularly where dimensions may be socially or causally related. Qualitative research and quantitative studies focusing on structural processes (e.g., discriminatory laws and policies, the unequal allocation of resources, or the deliberate siting of environmental hazards in particular communities)^47^ are needed to better understand the mechanisms underlying the inequalities documented here.

Furthermore, our findings relate to young people born in the UK in the early 2000s, and thus may not generalise to other UK youth (e.g., migrants or asylum seekers) (**Appendix 1, pp36-37**), or to other populations, generations, and contexts. Other intersecting systems of oppression (e.g., ableism, ageism, and religious discrimination) and social identities and positions (e.g., gender identity, alternative indicators of childhood socioeconomic circumstances, and current socioeconomic position) were not examined, and future studies may shed light on these.

Our study shows substantial social inequalities across multiple mental health outcomes among young people at the intersection of sexual identity, birth sex, childhood poverty, and ethnic identities/backgrounds. Although some intersectional positions experienced mental health outcomes that differed from additive expectations, most inequalities reflected the cumulative effects of multiple dimensions of social (dis)advantage. Health practitioners and policymakers can use this evidence to inform efforts to promote mental health equity. Our results demonstrate the value of examining multiple mental health outcomes through an intersectional lens, while also showing that commonly used measures may not function identically across social groups. Together, these findings underline the importance of identifying and addressing both the social conditions that generate mental health inequalities and the methodological assumptions that shape how those inequalities are measured and understood.

## Contributors

DMA conceptualised the study with contributions from all authors. DMA curated the data and conducted all statistical analyses. TS accessed and verified the data and code. All authors contributed to interpretation of results. DMA wrote the original draft and all authors reviewed and edited the manuscript. All authors had full access to all the data in the study and had final responsibility for the decision to submit for publication.

## Data sharing statement

Data from the Millennium Cohort Study is available at no-cost to bona fide researchers from the UK Data Service at http://doi.org/10.5255/UKDA-Series-2000031. Code for the data management and analyses is available in https://github.com/dmorenoagostino/intersectymh-mcs8. Parts of the analytical code were adapted from code from Evans and colleagues (https://doi.org/10.1016/j.ssmph.2024.101664).

## Data Availability

Data from the Millennium Cohort Study is available at no-cost to bona fide researchers from the UK Data Service at http://doi.org/10.5255/UKDA-Series-2000031.

## Acknowledgements

Funding for this study was provided by the Wellcome Trust [304283/Z/23/Z, 309118/Z/24/Z]. CRE is part supported by the UK Economic and Social Research Council (ESRC) [ES/X011313/1]. HLF is part supported by the UK ESRC Centre for Society and Mental Health at King’s College London [ES/S012567/1; UKRI861].

We would like to thank all the members of the ‘Making the margins visible’ Young People Advisory Group (YPAG) (including Adriana Vacaru, Akshay Gohil, Aleks Plewik, Alex Adams, Caitlin Brookes, Hemlata Pant, Jodayne Corrodus, Usayd Abid, and Yusra Nawaz, who consented to be named) for the input they have continuously provided to shape this study and the entire project, and to Hanna Jones and Evangelina Asiedu-Addo from McPin Foundation for facilitating the YPAG involvement. We would also like to thank all the participants of the Millennium Cohort Study for continuing to generously give up their time, and to the study team for their efforts in collecting and managing the data.

AI (M365 Copilot) was used to improve the readability of the manuscript and improve the code used for analyses.

## References

1. Tian J, Yan N, Hu X, et al. Global burden of mental disorders among adolescents and young adults, 1990-2021: a systematic analysis of the Global Burden of Diseases Study 2021. Gen Psychiatr 2025; 38(6): e102278.

2. Uhlhaas PJ, Davey CG, Mehta UM, et al. Towards a youth mental health paradigm: a perspective and roadmap. Mol Psychiatry 2023; 28(8): 3171–81.

3. Wang Z, Dou Y, Yang X, et al. Global, regional, and national burden of mental disorders among adolescents and young adults, 1990-2021: a systematic analysis for the Global Burden of Disease Study 2021. Translational psychiatry 2025; 15(1): 397.

4. Liubertiene G, Sloman A, Morris S, et al. Common mental health conditions. Adult Psychiatric Morbidity Survey: Survey of Mental Health and Wellbeing, England, 2023/4: NHS England; 2025.

5. Becares L, Kneale D. Inequalities in mental health, self-rated health, and social support among sexual minority young adults during the COVID-19 pandemic: analyses from the UK Millennium Cohort Study. Soc Psychiatry Psychiatr Epidemiol 2022; 57(10): 1979–86.

6. Moreno-Agostino D, Woodhead C, Ploubidis GB, Das-Munshi J. A quantitative approach to the intersectional study of mental health inequalities during the COVID-19 pandemic in UK young adults. Soc Psychiatry Psychiatr Epidemiol 2024; 59(3): 417–29.

7. Patalay P, Fitzsimons E. Development and predictors of mental ill-health and wellbeing from childhood to adolescence. Soc Psychiatry Psychiatr Epidemiol 2018; 53(12): 1311–23.

8. Irish M, Solmi F, Mars B, et al. Depression and self-harm from adolescence to young adulthood in sexual minorities compared with heterosexuals in the UK: a population-based cohort study. Lancet Child Adolesc Health 2019; 3(2): 91–8.

9. Chua YW, Schluter D, Pearce A, Sharp H, Taylor-Robinson D. Socioeconomic inequalities in mental health difficulties over childhood: a longitudinal sex-stratified analysis using the UK Millennium Cohort Study. Soc Sci Med 2025; 378: 118159.

10. Hazell M, Thornton E, Haghparast-Bidgoli H, Patalay P. Socio-economic inequalities in adolescent mental health in the UK: Multiple socio-economic indicators and reporter effects. Ssm-Ment Health 2022; 2.

11. Straatmann VS, Lai E, Lange T, et al. How do early-life factors explain social inequalities in adolescent mental health? Findings from the UK Millennium Cohort Study. J Epidemiol Community Health 2019; 73(11): 1049–60.

12. Moreno-Agostino D, Ploubidis GB, Das-Munshi J. COVID-19, economic downturn, and long-term trajectories of population mental health: Evidence from two nationally representative British birth cohorts at the intersection of gender and socioeconomic position. Soc Sci Med 2026; 389: 118830.

13. Terhaag S, Fitzsimons E, Daraganova G, Patalay P. Sex, ethnic and socioeconomic inequalities and trajectories in child and adolescent mental health in Australia and the UK: findings from national prospective longitudinal studies. J Child Psychol Psychiatry 2021; 62(10): 1255–67.

14. Crenshaw K. Demarginalizing the intersection of race and sex: A Black feminist critique of antidiscrimination doctrine, feminist theory, and antiracist politics. The University of Chicago Legal Forum 1989: 139–67.

15. Combahee River Collective. The Combahee River Collective Statement. 1977.

16. Bowleg L. A Framework for Applied Intersectionality Research (FAIR): Reframing Intersectionality as a Tool to Advance Health Equity and Social Justice Action, Not Just Empirical Research. Annual review of public health 2026; 47(1): 369–92.

17. Bowleg L, Bauer G. Invited Reflection: Quantifying Intersectionality. Psychol Women Quart 2016; 40(3): 337–41.

18. Fagrell Trygg N, Gustafsson PE, Mansdotter A. Languishing in the crossroad? A scoping review of intersectional inequalities in mental health. Int J Equity Health 2019; 18(1): 115.

19. Rohrer JM, Paulewicz B. Rethinking measurement invariance causally. Curr Res Ecol Soc Ps 2025; 9.

20. Patalay P, Fitzsimons E. Psychological distress, self-harm and attempted suicide in UK 17-year olds: prevalence and sociodemographic inequalities. The British journal of psychiatry: the journal of mental science 2021; 219(2): 437–9.

21. Connelly R, Platt L. Cohort profile: UK Millennium Cohort Study (MCS). Int J Epidemiol 2014; 43(6): 1719–25.

22. Joshi H, Fitzsimons E. The UK Millennium Cohort Study: the making of a multi-purpose resource for social science and policy in the UK. Longitudinal and Life Course Studies 2016; 7(4): 409–30.

23. Brown M, Fitzsimons E, Haselden L, et al. Millennium Cohort Study Sweep 8: Age 23 Survey User Guide (Version 2). London: UCL Centre for Longitudinal Studies; 2026.

24. Kessler RC, Green JG, Gruber MJ, et al. Screening for serious mental illness in the general population with the K6 screening scale: results from the WHO World Mental Health (WMH) survey initiative. International journal of methods in psychiatric research 2010; 19 Suppl 1(Suppl 1): 4–22.

25. Prochaska JJ, Sung HY, Max W, Shi Y, Ong M. Validity study of the K6 scale as a measure of moderate mental distress based on mental health treatment need and utilization. International journal of methods in psychiatric research 2012; 21(2): 88–97.

26. Stewart-Brown S, Tennant A, Tennant R, Platt S, Parkinson J, Weich S. Internal construct validity of the Warwick-Edinburgh Mental Well-being Scale (WEMWBS): a Rasch analysis using data from the Scottish Health Education Population Survey. Health Qual Life Outcomes 2009; 7: 15.

27. McAra L, McVie S. Youth crime and justice: Key messages from the Edinburgh Study of Youth Transitions and Crime. Criminol Crim Justic 2010; 10(2): 179–209.

28. McCall L. The complexity of intersectionality. Signs 2005; 30(3): 1771–800.

29. Asparouhov T, Muthén B. Multiple-Group Factor Analysis Alignment. Structural Equation Modeling-a Multidisciplinary Journal 2014; 21(4): 495–508.

30. Luong R, Flake JK. Measurement invariance testing using confirmatory factor analysis and alignment optimization: A tutorial for transparent analysis planning and reporting. Psychol Methods 2023; 28(4): 905–24.

31. Evans CR, Williams DR, Onnela JP, Subramanian SV. A multilevel approach to modeling health inequalities at the intersection of multiple social identities. Soc Sci Med 2018; 203: 64–73.

32. Evans CR, Leckie G, Subramanian SV, Bell A, Merlo J. A tutorial for conducting intersectional multilevel analysis of individual heterogeneity and discriminatory accuracy (MAIHDA). SSM Popul Health 2024; 26: 101664.

33. Tomova GD, Silverwood RJ, Tennant PWG, Wright L. How can the use of different modes of survey data collection introduce bias? An introduction to mode effects using directed acyclic graphs (DAGs). Am J Epidemiol 2026; 195(5): 1406–15.

34. StataCorp. Stata Statistical Software: Release 19. College Station, TX: StataCorp LLC; 2025.

35. Muthén LK, Muthén BO. Mplus User’s Guide. Eighth Edition ed. Los Angeles, CA:; 1998–2017.

36. Else-Quest NM, Hyde JS. Intersectionality in Quantitative Psychological Research: II. Methods and Techniques. Psychol Women Quart 2016; 40(3): 319–36.

37. Black L, Panayiotou M, Humphrey N. Estimating adolescent mental health in the general population: current challenges and opportunities. Lancet Psychiatry 2025; 12(2): 153–60.

38. Kelly D, Fitzsimons E. Mental health: Initial findings from the Millennium Cohort Study at Age 23. London: UCL Centre for Longitudinal Studies, 2026.

39. Wittgens C, Fischer MM, Buspavanich P, Theobald S, Schweizer K, Trautmann S. Mental health in people with minority sexual orientations: A meta-analysis of population-based studies. Acta Psychiatr Scand 2022; 145(4): 357–72.

40. Flynn SS, Touhey S, Sullivan TR, Mereish EH. Queer and Transgender Joy: A Daily Diary Qualitative Study of Positive Identity Factors among Sexual and Gender Minority Adolescents. Psychol Sex Orientat Gend Divers 2024.

41. Campbell OLK, Bann D, Patalay P. The gender gap in adolescent mental health: A cross-national investigation of 566,829 adolescents across 73 countries. SSM Popul Health 2021; 13: 100742.

42. Nazroo J. Race/ethnic inequalities in health: moving beyond confusion to focus on fundamental causes. Oxford Open Econ 2024; 3: i563–i76.

43. Ahmad G, McManus S, Cooper C, Hatch SL, Das-Munshi J. Prevalence of common mental disorders and treatment receipt for people from ethnic minority backgrounds in England: repeated cross-sectional surveys of the general population in 2007 and 2014. The British journal of psychiatry: the journal of mental science 2022; 221(3): 520–7.

44. Kirkbride JB, Anglin DM, Colman I, et al. The social determinants of mental health and disorder: evidence, prevention and recommendations. World psychiatry: official journal of the World Psychiatric Association 2024; 23(1): 58–90.

45. Mokhwelepa LW, Sumbane GO. Men’s Mental Health Matters: The Impact of Traditional Masculinity Norms on Men’s Willingness to Seek Mental Health Support; a Systematic Review of Literature. Am J Mens Health 2025; 19(3): 15579883251321670.

46. McGarrity LA. Socioeconomic status as context for minority stress and health disparities among lesbian, gay, and bisexual individuals. Psychology of Sexual Orientation and Gender Diversity 2014; 1(4): 383.

47. Homan P, Brown TH, King B. Structural Intersectionality as a New Direction for Health Disparities Research. J Health Soc Behav 2021; 62(3): 350–70.

