## Appendix for "Intersectional inequalities in youth mental health measurement and outcomes: population-representative evidence from the UK Millennium Cohort Study": Appendix 1.docx

### 1. STROBE statement

|  | | Item No | Recommendation | Page  No |
| --- | --- | --- | --- | --- |
| **Title and abstract** | | 1 | (*a*) Indicate the study’s design with a commonly used term in the title or the abstract | 2 |
|  |  |  | (*b*) Provide in the abstract an informative and balanced summary of what was done and what was found | 2 |
| Introduction | | | | |
| Background/rationale | | 2 | Explain the scientific background and rationale for the investigation being reported | 3 |
| Objectives | | 3 | State specific objectives, including any prespecified hypotheses | 3 |
| Methods | | | | |
| Study design | | 4 | Present key elements of study design early in the paper | 3-4 |
| Setting | | 5 | Describe the setting, locations, and relevant dates, including periods of recruitment, exposure, follow-up, and data collection | 3-4 |
| Participants | | 6 | (*a*) *Cohort study*—Give the eligibility criteria, and the sources and methods of selection of participants. Describe methods of follow-up  *Case-control study*—Give the eligibility criteria, and the sources and methods of case ascertainment and control selection. Give the rationale for the choice of cases and controls  *Cross-sectional study*—Give the eligibility criteria, and the sources and methods of selection of participants | 3-4 |
|  |  |  | (*b*) *Cohort study*—For matched studies, give matching criteria and number of exposed and unexposed  *Case-control study*—For matched studies, give matching criteria and the number of controls per case |  |
| Variables | | 7 | Clearly define all outcomes, exposures, predictors, potential confounders, and effect modifiers. Give diagnostic criteria, if applicable | 4, Appendix 1 |
| Data sources/ measurement | | 8* | For each variable of interest, give sources of data and details of methods of assessment (measurement). Describe comparability of assessment methods if there is more than one group | 4, Appendix 1 |
| Bias | | 9 | Describe any efforts to address potential sources of bias | 5 |
| Study size | | 10 | Explain how the study size was arrived at | 3-4 |
| Quantitative variables | | 11 | Explain how quantitative variables were handled in the analyses. If applicable, describe which groupings were chosen and why | 4, Appendix 1 |
| Statistical methods | | 12 | (*a*) Describe all statistical methods, including those used to control for confounding | 4-5, Appendix 1 |
|  |  |  | (*b*) Describe any methods used to examine subgroups and interactions | 4-5, Appendix 1 |
|  |  |  | (*c*) Explain how missing data were addressed | 4-5, Appendix 1 |
|  |  |  | (*d*) *Cohort study*—If applicable, explain how loss to follow-up was addressed  *Case-control study*—If applicable, explain how matching of cases and controls was addressed  *Cross-sectional study*—If applicable, describe analytical methods taking account of sampling strategy | 4-5, Appendix 1 |
|  |  |  | (*e*) Describe any sensitivity analyses | 5, Appendix 1 |
| Results | | | | |
| Participants | 13* | (a) Report numbers of individuals at each stage of study—eg numbers potentially eligible, examined for eligibility, confirmed eligible, included in the study, completing follow-up, and analysed | | 6 |
|  |  | (b) Give reasons for non-participation at each stage | | 6 |
|  |  | (c) Consider use of a flow diagram | |  |
| Descriptive data | 14* | (a) Give characteristics of study participants (eg demographic, clinical, social) and information on exposures and potential confounders | | 6, Table 1, Appendix 1 |
|  |  | (b) Indicate number of participants with missing data for each variable of interest | | Table 1, Appendix 1 |
|  |  | (c) *Cohort study*—Summarise follow-up time (eg, average and total amount) | |  |
| Outcome data | 15* | *Cohort study*—Report numbers of outcome events or summary measures over time | |  |
|  |  | *Case-control study—*Report numbers in each exposure category, or summary measures of exposure | |  |
|  |  | *Cross-sectional study—*Report numbers of outcome events or summary measures | | Table 1, Appendix 1 |
| Main results | 16 | (*a*) Give unadjusted estimates and, if applicable, confounder-adjusted estimates and their precision (eg, 95% confidence interval). Make clear which confounders were adjusted for and why they were included | | 6-7, Figure 1-3, Appendix 1, Appendix 2 |
|  |  | (*b*) Report category boundaries when continuous variables were categorized | |  |
|  |  | (*c*) If relevant, consider translating estimates of relative risk into absolute risk for a meaningful time period | |  |
| Other analyses | 17 | Report other analyses done—eg analyses of subgroups and interactions, and sensitivity analyses | | 8, Appendix 1, Appendix 2 |
| Discussion | | | | |
| Key results | 18 | Summarise key results with reference to study objectives | | 8-9 |
| Limitations | 19 | Discuss limitations of the study, taking into account sources of potential bias or imprecision. Discuss both direction and magnitude of any potential bias | | 10 |
| Interpretation | 20 | Give a cautious overall interpretation of results considering objectives, limitations, multiplicity of analyses, results from similar studies, and other relevant evidence | | 10 |
| Generalisability | 21 | Discuss the generalisability (external validity) of the study results | | 10, Appendix 1 |
| Other information | | | | |
| Funding | 22 | Give the source of funding and the role of the funders for the present study and, if applicable, for the original study on which the present article is based | | 11 |

*Give information separately for cases and controls in case-control studies and, if applicable, for exposed and unexposed groups in cohort and cross-sectional studies.

**Note:** An Explanation and Elaboration article discusses each checklist item and gives methodological background and published examples of transparent reporting. The STROBE checklist is best used in conjunction with this article (freely available on the Web sites of PLoS Medicine at http://www.plosmedicine.org/, Annals of Internal Medicine at http://www.annals.org/, and Epidemiology at http://www.epidem.com/). Information on the STROBE Initiative is available at www.strobe-statement.org.

### 2. Additional details on outcome measures

The following outcome measures, obtained from data self-reported by the participants at the age 23 sweep/survey wave, were used:

**Psychological distress** was measured with the ‘Kessler-6’ (K6) questionnaire.^1^ The K6 questionnaire includes six questions on the frequency the respondent experienced different mental ill-health symptoms in the past 30 days: depressed, hopeless, restless or fidgety, that everything was an effort, worthless, and nervous. Response options range from 0 (“None of the time”) to 4 (“All of the time”). Scores were derived as the sum of the responses to the six questions, ranging between 0 (minimum distress) to 24 (maximum distress). In workshops with the Young People Advisory Group (YPAG) involved in this project, participants identified the fifth question (“How often did you feel worthless?” as the most likely one to be interpreted similarly across young people from different social positions and identities.

**‘Caseness’ (or probable serious psychological distress)** was operationalised as a Kessler 6 score of 13 or higher, a threshold that has been identified to provide good balance between sensitivity and specificity to classify individuals with mental health treatment needs (e.g., AUC = 0.82-0.87).^2,3^

**Psychological well-being** was measured with the ‘Short Warwick-Edinburgh Mental Well-Being Scale’ (SWEMWBS).^4^ The SWEMWBS includes seven questions on the frequency the respondent has felt different positive experiences in the last two weeks: optimistic about the future, useful, relaxed, dealing with problems well, thinking clearly, close to other people, and able to make up their own mind about things. Response options range from 1 (“None of the time”) to 5 (“All of the time”). Scores were derived as the sum of the responses to the seven questions, ranging between 1 (minimum wellbeing) to 35 (maximum wellbeing). In workshops with the YPAG involved in this project, participants identified the third statement (“I’ve been feeling relaxed”) as the most likely one to be interpreted similarly across young people from different social positions and identities.

**Any self-harm in the last 12 months** was measured by a series of six questions, based on the Edinburgh Study of Youth Transitions,^5^ on whether, during the last year, the respondent had hurt themself on purpose in any of the following ways: cut or stabbed, burned, or bruised or pinched themself, taken an overdose of tablets, pulled out their hair, or hurt themself in some other way. Response options were two: “Yes” or “No”. A binary variable indicating whether the cohort member had responded “Yes” to any of the previous questions was created, reflecting whether they had hurt themself on purpose in any way during the last year. For sensitivity analyses, an additional variable was derived as the total number of self-harm behaviours reported, ranging from 0 (none) to 6 (all).

**Lifetime suicide attempts** were measured by a single question on whether the respondent has ever hurt themself on purpose in an attempt to end their life, with two response options: “Yes” or “No”. Responses to the same question at the age 17 sweep/survey wave were used to inform values in this outcome. Where an affirmative response had been provided at the age 17 sweep but not at age 23 sweep (either due to negative response or missing data, including refusal), the response at age 17 sweep was retained.

### 3. Additional details on variables used to define intersectional strata/positions

Consistent with an inter-categorical approach to intersectionality,^6^ we provisionally used data on four social identities and positions as proxies for the systems of oppression under study:

**Sex assigned at birth** was used as a proxy for sexism. We used data obtained at the age 9 months sweep/wave, coded by the interviewer and asked to the interviewed parent ‘if not obvious’ (as per the instructions in the computer assisted personal interviewing script), with options “Male”, and “Female”. This was supplemented by information from subsequent sweeps in cases where that information was missing.

**Ethnicity** was used as a proxy for racism. We prioritised self-designated ethnic identity as provided by the cohort member at the age 14 sweep as a response to the question “What is your ethnic group?”. Response options to that question included: “White” (with further options: “English/Welsh/Scottish/Northern Irish/British”, “Irish”, “Gypsy or Irish Traveller”, “Any other White background”), “Mixed/multiple ethnic groups (with further options: “White and Black Caribbean”, “White and Black African”, “White and Asian”, “Any other mixed/multiple ethnic background”), “Asian/Asian British” (with further options: “Indian”, “Pakistani”, “Bangladeshi”, “Chinese”, “Any other Asian background”), “Black/African/Caribbean/Black British” (with further options: “African”, “Caribbean”, “Other Black/African/Caribbean background”), and “Other ethnic group” (with further options: “Arab”, “Other ethnic group”). The further response options slightly varied by UK country, with the above corresponding to the options for interviews in England. In cases where self-designated information was missing, we complemented it, first, by parent-reported information at previous waves, in response to the question “Which of the groups on this card do you regard [cohort child] as belonging to?”, with similar response options. If information on ethnicity was still missing, we further used data on self-designated identity provided by the natural parents, if available. Due to data constraints, we grouped together smaller ethnic groups into the following six categories, aiming to keep as much granularity as possible and with input from the YPAG involved in this project: White (including all White groups), Mixed (including all Mixed groups), Indian, Pakistani/Bangladeshi (combined), Black (including all Black groups), and Other (including the “Chinese”, “Any other Asian background”, and “Other ethnic group” categories). Descriptive statistics at the more granular categories available in the data are provided in pages 17-22 in this Appendix.

**Sexual identity** was used as a proxy for heterosexism. Sexual identity was obtained from the question “Which of the following options best describes how you currently think of your sexuality?” collected at the latest sweep/wave (age 23). Although original response options included “Completely heterosexual/straight”, “Mainly heterosexual/straight”, “Bisexual”, “Mainly gay or lesbian”, “Completely gay or lesbian”, “Other”, a derived variable with the following categories was deposited in the UK Data Service as part of the End User License deposit: “Heterosexual”, “Gay or lesbian”, “Bisexual”, “Pansexual”, “Asexual (including demisexual)”, “Queer”, “All other sexualities”.^7^ This meant we were no longer able to distinguish individuals who may have reported identifying as “completely” vs “mainly heterosexual/straight”, which previous research has suggested may have differential outcomes.^8^ Due to data constraints, all categories except “Heterosexual” were grouped together as a “Sexual minority” category. In cases where this information was missing, it was supplemented by a similar question asked at the previous sweep (age 17). Although this can blur inequalities across some of the categories within the “Sexual minority” group, previous research has shown that, although to different degrees, all these groups experience worse mental health and ill-health outcomes than the “Heterosexual” group.^8^ Descriptive statistics at the more granular categories available in the age 23 sweep data deposit are provided in pages 17-22 in this Appendix.

**Childhood poverty** was used as a measure of early-childhood socioeconomic deprivation and a proxy for classism. This measure was constructed using household-level income data collected at two time points: 9 months and 3 years. Socioeconomic inequalities in mental health have been shown to be firmly established by exposure to socioeconomic adversity in early life.^9,10^ At these two sweeps, the Millennium Cohort Study (MCS) team derived an indicator of household poverty based on OECD equivalised income, coded as below or above 60% of the national median income, consistent with standard UK definitions of relative poverty/low income. Using these sweep-specific indicators, we created a binary variable representing whether the cohort member had “ever” vs “never” lived in a household below the 60% median income threshold at any of the two time points. In cases where information at 9 months and 3 years was missing, data collected between ages 5 and 14 years was used. In those cases, if every available observation was consistently above or below the 60% of the national median income, that value was assigned to the childhood poverty binary indicator.

### 4. Additional details on the measurement invariance/equivalence testing rationale and approach

We used a measurement invariance/equivalence testing approach to address the first objective of the study (to investigate whether commonly used measures of mental ill-health and wellbeing were comparable across social identities, positions, and their intersections). Unbiased comparisons of outcomes across groups require that these are not due to differences in how these have been measured.^11^ This is often ignored in research involving multiple groups, and engagement with this in quantitative intersectional research is almost non-existent, despite calls for its application from at least a decade ago.^12^

We used a combination of two approaches that are suitable for empirically testing whether such differences exist across multiple groups when using multiple-indicator outcome measures such as the Kessler-6 and SWEMWBS: multiple-group confirmatory factor analysis (MG-CFA)^13^ and the alignment method or optimisation.^14^ Luong and Flake provide an excellent overview and comparison of both approaches.^15^

We first estimated unidimensional CFA models on the entire sample to examine the appropriateness of the factor structure. In line with existing guidelines,^16^ overall model fit for the unidimensional single-group CFA models was considered to be good if the comparative fit index (CFI) and Tucker-Lewis Index (TLI) values were higher than 0.950, the root mean square error of approximation (RMSEA) value was lower than 0.060, and the standardised root mean square residual (SRMR) was lower than 0.08. CFI/TLI > 0.900 and RMSEA < 0.080 were considered as acceptable.

Although the indicators in both the Kessler-6 and the SWEMWBS are ordinal, we treated them as continuous. Given the small sample sizes in many of the groups under study, this substantially reduced model complexity (one intercept per item rather than multiple thresholds) and helped avoid estimation problems arising from sparse response categories. However, to accommodate departures from normality, all models were estimated using robust maximum likelihood (MLR).

Alignment was used as a computationally efficient screening approach across numerous intersectional groups, with the more flexible MG-CFA approach subsequently used to examine and address non-invariance in greater detail where required.

#### 4.1. Alignment method: screening for approximate measurement invariance

To screen for potential departures from approximate measurement invariance, we used the alignment method. This method algorithmically attempts to reach a solution where most measurement parameters under study (in this case, loading and intercepts or thresholds) are approximately equal (i.e., that approximate measurement invariance holds).^15^ Following existing recommendations,^17^ measures were considered sufficiently comparable if no more than 25% of the measurement parameters were flagged as non-invariant (with α=0.001).

Due to small sample sizes at some intersectional strata, we implemented these models from more general to more granular scenarios. Hence, we first ran the alignment models by single variables (i.e., birth sex, ethnicity, sexual identity, and childhood poverty), and then by two-way interactions (e.g., birth sex by ethnicity). We excluded groups with less than 20 observations as their inclusion led to convergence issues or inappropriate solutions (e.g., negative variance estimates). Therefore, the most granular alignment stratification analysed was the three-way interaction of birth sex, sexual identity, and childhood poverty, as three-way interaction terms including ethnicity would have resulted in too many excluded groups (see eTable 10.2, page 24, in this Appendix).

#### 4.2. MG-CFA: detailed analysis of partial measurement invariance

In cases where approximate invariance did not hold based on the above thresholds, we used an MG-CFA approach. In this approach, models with increasing constraints (on the equality of measurement parameters across the groups) are estimated and compared to ascertain whether the corresponding loss in model fit resulting from the equality constraints is acceptable.

The estimated models are, typically:

1) A configural invariance model, testing that the configuration of factors and indicators reasonably holds across groups;
2) A metric/weak invariance model, testing that the loadings, representing the strength with which the different indicators relate to the factors, are the same across groups;
3) And a scalar/strong invariance model, testing that the intercepts or thresholds, representing the expected level in the indicator when the factor equals zero [intercept] or the point in the underlying response continuum in which the probability to provide two contiguous response options is the same [threshold], are the same across groups. This level of invariance is required for unbiased comparisons of the outcome means across groups.

Satisfactory model fit is typically considered to be achieved if the loss in fit between configural (with no equality constraints across intersectional positions/strata in loadings or thresholds) and metric invariance (equality constraints in loadings) models, and between metric and scalar invariance (equality constraints in loadings and thresholds) models is smaller than 0.015 for the root mean square error of approximation (RMSEA) and 0.010 for the comparative fit index (CFI).^18^

Partial measurement invariance can be achieved even when some measurement parameters are non-invariant, provided that a majority of parameters remain invariant across groups.^15^ This can be done by exploring the modification indices of the models with constraints to identify which specific parameters could be freed to improve fit. These parameters can be kept free while keeping the others invariant, assessing the resulting comparative fit. If this is satisfactory, based on the abovementioned thresholds for RMSEA and CFI loss, the measurement invariance testing can proceed.

We used MG-CFA models in all cases where the implementation of the alignment method showed that approximate measurement invariance did not hold, as well as further multiple-way interactions involving that single variable or interaction with sufficient number of observations per stratum (few strata with less than 20 observations).

Anchor/referent items for each measure (items that are assumed to be invariant across groups) were selected with input from the YPAG involved in this project, who helped identify which items they considered to be the most likely one to be interpreted similarly across young people from different social positions and identities in both the Kessler-6 (“How often did you feel worthless?”) and SWEMWBS (“I’ve been feeling relaxed”).

Factor scores were then derived from the most granular and most invariant models identified for each outcome as the maximum of the posterior distribution of the factor (also known as the regression method for factor score estimation) and used in the main analyses to account for identified measurement non-invariance.

Detailed results from the measurement invariance/equivalence testing models are provided in pages 23-27 in this Appendix.

### 5. Additional details on the I-MAIHDA modelling approach used

We used Intersectional Multilevel Analysis of Individual Heterogeneity and Discriminatory Accuracy (I-MAIHDA)^19^ models to test the hypothesis of social and intersectional inequalities in mental health and ill-health outcomes. These are multilevel models where individual observations are nested within intersectional positions or strata.

For continuous outcomes (the partially invariant factor scores and the observed sum scores of Kessler-6 and SWEMWBS), we used multilevel linear models. For binary outcomes (‘caseness’ or probable serious psychological distress, any self-harm, and any suicide attempt), we used multilevel logistic models. For count outcomes (number of self-harm behaviours), we used negative binomial and Poisson models.

First, intercepts-only (null) models (multilevel models including the corresponding outcome variable) were estimated to calculate the degree of clustering within intersectional strata, as measured by the proportion of total variance that is due to between-strata differences (the Variance Partition Coefficient, VPC_null_). VPC estimates range between 0 (indicating no differences between strata, with all variability taking place within strata) and 1 (indicating that all variance is due to differences between strata and that there is no variability within strata).

Subsequent models including the main (or additive) fixed effects of the variables defining the intersectional strata were then estimated. We calculated the proportional change in variance (PCV) as the difference in between-strata variance between the intercepts-only and main models, divided by the between-strata variance of the intercepts-only model. The PCV takes values between 0 (indicating no change, and therefore suggesting that none of the between-strata variance is due to the main or non-intersectional effects of each of the variables defining the intersectional strata) and 1 (indicating that there is no remaining between-strata variance after accounting for those main or additive effects). We partitioned the variance from linear and logistic I-MAIHDA models following existing I-MAIHDA guidelines.^19,20^ To partition the variance from binomial and Poisson models, we followed the indications in Leckie *et al*.^21^

To the extent that models indicated departures from additivity (i.e., departures from the general inequality patterns captured by the main effects, as suggested by PCV values lower than 1), we further investigated these by estimating the stratum-specific random/residual effects as the empirical Bayes (EB) predictions. We interpreted these as estimates of the intersectional effects at each intersectional stratum, with larger values (either positive or negative) representing a larger departure from the levels expected based on the main or additive effects of each of the variables defining the intersectional strata. Because we used intersectionality as a framework, lens, or paradigm throughout the study rather than as a “testable hypothesis”,^22^ lack of evidence of intersectional effects based on the terms established above was not interpreted as “lack of empirical proof” for intersectionality.

Regardless of the presence or absence of “significant” (different-than-zero) intersectional effects, we used the main models (i.e., models adjusted for the variables defining the intersectional strata) to generate predicted mean levels (for continuous outcomes), probabilities (for binary outcomes), and number of behaviours (for count outcomes) for each intersectional stratum to characterise inequalities across strata, since I-MAIHDA models can provide more accurate estimates than fixed-effects models even with small group sample sizes.^23^

### 6. Additional details on quantitative bias analysis (QBA) of mode effects

The age 23 Millennium Cohort Study (MCS) sweep (or data collection) used a sequential mixed-mode design: online survey, followed by face-to-face interviews for non-respondents, and a shortened online-only mop-up survey for remaining non-respondents (making up around 4.7% of the productive interviews).^7^

Mental health measures can be biased by “mode effects”, whereby responses to some questions can vary depending on the administration mode. Administration modes that provide more privacy, including computerised modes, tend to elicit more negative responses to sensitive questions, including questions on mental health.^24,25^

Because participants were not randomly allocated to different survey modes, mode effects can be compounded by mode selection, rendering the simple adjustment by the administration mode inappropriate to account for mode effects.^26^

Questions on mental health in the age 23 MCS sweep were administered as part of a self-completion questionnaire due to their sensitive nature,^7^ which may minimise the impact of mode effects. Nevertheless, we used a quantitative bias analysis approach to explore the potential impact of these effects on the results, focusing on the psychological distress (Kessler-6) and wellbeing (SWEMWBS) measures.

Our approach consisted of applying a “correction factor” or deviation to the partially invariant Kessler-6 and SWEMWBS factor scores among those who participated in face-to-face interviews to simulate what could have been expected had it been collected online. We were not able to find previous evidence on the expected mode effects in these measures across the administration modes used in MCS. Therefore, we explored the impact of different amounts of bias across six different scenarios.

In Scenarios 1 to 3, in-person respondents were assumed to homogeneously underreport distress and overreport wellbeing, and their scores were adjusted by +0.2, +0.4, and +0.4 standard deviation units for Kessler-6 and -0.2, -0.4, and -0.6 standard deviation units for SWEMWBS, respectively.

Scenarios 4–6 extended these assumptions by allowing mode effects to vary across groups, with larger adjustments applied to sexual minority participants, ethnic minority groups, and those who experienced childhood poverty. Scores in these groups were adjusted by +0.1, +0.2, and +0.3 standard deviation units for Kessler-6 and -0.1, -0.2, -0.3 standard deviation units for SWEMWBS, respectively, for each of these social identities and positions. Therefore, for instance, a face-to-face participant from an ethnic and sexual minority and who experienced childhood poverty would see their Kessler-6 score adjusted by +0.5 standard deviation units in Scenario 4 (+0.2 homogeneous correction factor, +0.1 sexual minority, +0.1 ethnic minority, +0.1 childhood poverty), and by +1.5 standard deviation units in Scenario 6 (+0.6 homogeneous correction factor, +0.3 sexual minority, +0.3 ethnic minority, +0.3 childhood poverty).

These scenarios reflect the possibility that social desirability, stigma, or reduced privacy may differentially influence reporting. However, as it can be seen from the example above, the relatively arbitrary “correction factors” implemented can result in quite extreme simulated values. Therefore, the aim was for these analyses to serve as “tipping point analyses” (how large should bias be to substantially alter the findings) rather than to provide corrected estimates.

### 7. Additional details on the lived experience involvement

This study, and the entire project that it is part of (“Making the margins visible”), has been shaped by the involvement of a Young People Advisory Group (YPAG) comprised of multiple young people from diverse intersectional positions and with lived experience of mental ill-health and/or social discrimination.

Regular workshops were held with the YPAG, where key aspects of the research project as a whole and of this study more specifically were discussed. This included, for example, discussions around what language to use (and to avoid) when talking about different social identities and positions, and what outcomes and measures to prioritise in this and subsequent studies.

Once analyses started, discussions were held with the wider YPAG on the preferences for how to group ethnic identity/background categories in the face of small sample sizes at specific intersectional positions. Their input directly translated into this study keeping the combined Pakistani and Bangladeshi group separate from the Indian group rather than grouping them together as is often done.

Input from the wider YPAG was also embedded into the measurement invariance/equivalence testing process, where their insights into which Kessler-6 and SWEMWBS items were more likely to be interpreted similarly by young people from different backgrounds directly informed which items were used as anchor items in the partial measurement invariance models.

A discussion with the wider YPAG was also carried out to identify which findings they considered to be the key ones, what additional context may be needed to adequately interpret them considering their lived experience, and how to frame the interpretation of the results to avoid unintentional harm to groups within the young population and to minimise misuse of the findings. We also discussed what section of the young population may not be represented in the study, with input from these discussions directly feeding into the section “Silences and absences: who is not (sufficiently) represented in this study”, in this Appendix (pages 36-37).

Smaller groups of YPAG members directly contributed to the preregistered protocol (<https://doi.org/10.17605/OSF.IO/U89GR>), which includes those YPAG contributors who desired to be named, and to reviewing and editing the manuscript draft and appendix before submission.

Across all these different engagement forms, members of the YPAG were offered training in notions of population data and analysis, and were remunerated for their time in line with National Institute for Health and Care Research (NIHR) guidance (<https://www.nihr.ac.uk/about-us/who-we-are/policies-and-guidelines/payment-guidance-researchers-and-professionals>).

### 8. Full list of deviations from preregistered study protocol

| **Preregistered approach** | **Deviation** | **Reason for deviation** |
| --- | --- | --- |
| On measurement of self-harm: “Factor scores reflecting self-harm propensity will be derived using models optimised to minimise measurement invariance across groups” | We did not derive these measures of self-harm propensity. | Measurement models including a latent factor representing self-harm propensity were unstable, largely due to empty cells across many of the individual self-harm indicators. |
| On operationalisation of ethnic identity/background: “In cases where self-designated information was missing, this will be complemented by parent-reported information at previous waves, in response to the question “Which of the groups on this card do you regard [cohort child] as belonging to?”, with similar response options.” | We further complemented this information with information on the natural parents’ ethnicity if this was available. | Maximise the use of available data to minimise missing data. |
| On operationalisation of sexual identity: “due to the aims of the study (see “Gender” paragraph), we expect we will need to dichotomise this variable into “Completely heterosexual/straight” and “Sexual minority”, with the latter including all the categories other than “Completely heterosexual/straight”.” | “Mainly heterosexual/straight” was included as part of the “Heterosexual” group. | A derived variable containing recoded groups rather than the original response options is provided in the End User License data deposit. This derived variable does not disaggregate between “completely” and “mainly” heterosexual participants, including them both in the “Heterosexual” group. |
| On item missingness: “missing data will be imputed using data predictive of the missingness and the values of the missing data, collected at the latest and previous sweeps/waves, using Multiple Imputation by Chained Equations (MICE).” | We did not use multiple imputation to deal with item missingness. | Outcome-specific missingness ranged from 3.0% to 3.5%. Limitations in the implementation of multiple imputation within I-MAIHDA models (including difficulties to ensure congeniality between imputation and analysis models), and the use of non-response weights to account for unit missingness, led us to discard the use of multiple imputation. |
| On the implementation of measurement invariance/equivalence testing: “unidimensional latent factors will be measured by the respective categorical indicators” | We modelled the indicators as continuous. | This substantially reduced model complexity (one intercept per item rather than multiple thresholds) and helped avoid estimation problems arising from sparse response categories. We accommodated departures from normality by using robust maximum likelihood (MLR) estimation. |

### 9. Descriptive statistics by granular categories (before recoding for analyses) and visualisations

**eTable 9.1** shows the distribution of social identities and positions and outcomes estimates (weighted results) before recoding/grouping categories up. The number of missing ethnic identity observations is larger than in the main analyses because using the information on the ethnic identity of their natural parents’ allowed us to assign some individuals to broader categories (e.g., Mixed) but not necessarily to the specific subcategory. Similarly, information on sexual identity in **eTable 9.1** is based on the information at age 23, not augmented with information from any previous waves as the granular categories were different.

A visualisation of the estimates in each of the more granular levels of ethnicity and sexual identity is provided in **eFigure 9.1** (ethnicity) and **eFigure 9.2** (sexual identity).

A visualisation of the estimates in each of the categories as used in the analyses is provided in **eFigure 9.3**.

##### eTable 9.1. Distribution of social identities and positions and outcome estimates before recoding for analyses, weighted results.

|  |  |  |  | **Psychological distress (Kessler-6)** | | **Caseness  (Kessler-6 ≥ 13)** | | **Psychological wellbeing  (SWEMWBS)** | | **Any self-harm  in past 12 months** | | **Self-harm (number  of behaviours)**** | | **Lifetime attempted suicide** | |
| --- | --- | --- | --- | --- | --- | --- | --- | --- | --- | --- | --- | --- | --- | --- | --- |
|  | **N obs** | **Unweighted %** | **Weighted % *** | **N obs** | **Mean (95% CI) *** | **N obs** | **Prevalence (95% CI) *** | **N obs** | **Mean (95% CI) *** | **N obs** | **Prevalence (95% CI) *** | **N obs** | **Mean (95% CI) *** | **N obs** | **Prevalence (95% CI) *** |
| **Overall** | 9675 |  |  | 9349 | 8.4 (8.2, 8.6) | 9350 | 21.3 (20.1, 22.5) | 8905 | 23.3 (23.2, 23.4) | 8900 | 20.3 (19.1, 21.6) | 8881 | 0.4 (0.4, 0.4) | 8943 | 13.1 (12.1, 14.2) |
| **Sex assigned at birth** |  |  |  |  |  |  |  |  |  |  |  |  |  |  |  |
| Female | 5240 | 54.2% | 50.9% | 5066 | 9.2 (9.0, 9.5) | 5067 | 25.5 (23.7, 27.3) | 4847 | 22.9 (22.7, 23.1) | 4842 | 24.3 (22.5, 26.2) | 4829 | 0.5 (0.4, 0.5) | 4871 | 16.9 (15.3, 18.6) |
| Male | 4434 | 45.8% | 49.1% | 4282 | 7.5 (7.3, 7.8) | 4282 | 16.9 (15.4, 18.6) | 4057 | 23.8 (23.6, 23.9) | 4057 | 16.1 (14.5, 17.8) | 4051 | 0.3 (0.3, 0.3) | 4071 | 9.1 (8.0, 10.4) |
| *Missing* | *1* |  |  |  |  |  |  |  |  |  |  |  |  |  |  |
| **Ethnic identity/background** |  |  |  |  |  |  |  |  |  |  |  |  |  |  |  |
| White | 7708 | 79.7% | 85.1% | 7474 | 8.3 (8.1, 8.5) | 7475 | 20.7 (19.4, 22.0) | 7109 | 23.4 (23.3, 23.6) | 7099 | 20.6 (19.3, 22.0) | 7084 | 0.4 (0.4, 0.4) | 7132 | 13.4 (12.3, 14.5) |
| Mixed White & Black Caribbean | 122 | 1.3% | 1.2% | 119 | 9.2 (8.1, 10.3) | 119 | 26.7 (17.6, 38.3) | 112 | 22.4 (21.1, 23.7) | 112 | 14.3 (8.2, 23.8) | 112 | 0.4 (0.1, 0.6) | 113 | 13.5 (7.8, 22.4) |
| Mixed White & Black African | 47 | 0.5% | 0.5% | 44 | 10.0 (7.9, 12.1) | 44 | 28.3 (13.1, 50.6) | 43 | 21.6 (19.9, 23.2) | 43 | 14.1 (6.5, 28.0) | 43 | 0.3 (0.1, 0.5) | 44 | 22.6 (9.2, 45.8) |
| Mixed White & Asian | 129 | 1.3% | 1.2% | 122 | 10.0 (8.8, 11.2) | 122 | 30.5 (21.4, 41.4) | 120 | 22.9 (22.0, 23.9) | 121 | 27.6 (18.9, 38.5) | 120 | 0.6 (0.3, 0.9) | 121 | 14.3 (8.2, 23.8) |
| Other Mixed | 121 | 1.3% | 1.5% | 116 | 9.5 (7.8, 11.3) | 116 | 34.1 (19.1, 53.3) | 112 | 23.0 (21.9, 24.1) | 112 | 38.0 (22.2, 56.9) | 112 | 1.0 (0.4, 1.6) | 112 | 16.3 (8.9, 27.8) |
| Indian | 268 | 2.8% | 1.8% | 256 | 8.8 (7.9, 9.8) | 256 | 25.7 (18.8, 34.2) | 246 | 22.9 (22.1, 23.7) | 244 | 16.4 (11.0, 23.7) | 244 | 0.3 (0.2, 0.4) | 246 | 12.5 (7.5, 20.1) |
| Pakistani | 507 | 5.2% | 2.9% | 482 | 8.1 (7.3, 8.8) | 482 | 19.9 (15.5, 25.2) | 461 | 23.1 (22.5, 23.7) | 464 | 12.2 (8.5, 17.1) | 462 | 0.2 (0.1, 0.3) | 467 | 4.6 (3.1, 6.7) |
| Bangladeshi | 205 | 2.1% | 0.9% | 190 | 8.5 (7.6, 9.5) | 190 | 17.3 (11.7, 24.8) | 183 | 22.3 (21.3, 23.2) | 186 | 11.1 (6.4, 18.4) | 185 | 0.3 (0.1, 0.5) | 186 | 6.1 (2.8, 12.8) |
| Other Asian | 111 | 1.1% | 0.8% | 110 | 8.1 (6.9, 9.4) | 110 | 15.6 (8.6, 26.7) | 104 | 22.8 (21.6, 24.0) | 104 | 15.0 (8.3, 25.5) | 104 | 0.3 (0.1, 0.5) | 105 | 9.1 (4.5, 17.8) |
| Black African | 196 | 2.0% | 1.7% | 190 | 8.2 (6.8, 9.5) | 190 | 19.1 (11.6, 29.9) | 182 | 23.2 (22.2, 24.2) | 180 | 11.9 (7.0, 19.3) | 180 | 0.2 (0.1, 0.2) | 180 | 14.7 (6.1, 31.3) |
| Black Caribbean | 105 | 1.1% | 0.9% | 97 | 9.8 (8.6, 11.0) | 97 | 25.3 (16.0, 37.7) | 92 | 22.8 (21.8, 23.8) | 94 | 15.7 (8.0, 28.5) | 94 | 0.3 (0.0, 0.7) | 96 | 17.1 (9.0, 30.2) |
| Other Black | 27 | 0.3% | 0.2% | 27 | 9.4 (7.3, 11.6) | 27 | 12.6 (4.2, 32.5) | 27 | 22.5 (20.9, 24.2) | 26 | 6.8 (1.6, 24.5) | 26 | 0.1 (0.0, 0.2) | 26 | 6.6 (1.5, 24.1) |
| Arab | 79 | 0.8% | 0.4% | 75 | 8.9 (7.0, 10.9) | 75 | 30.5 (17.7, 47.2) | 71 | 21.2 (19.8, 22.6) | 72 | 24.8 (13.4, 41.2) | 72 | 0.5 (0.2, 0.9) | 71 | 14.7 (5.9, 32.1) |
| Other | 28 | 0.3% | 0.4% | 27 | 10.2 (8.4, 12.0) | 27 | 26.5 (10.5, 52.7) | 25 | 22.4 (20.4, 24.3) | 25 | 37.9 (13.1, 71.2) | 25 | 0.9 (0.0, 1.9) | 26 | 15.7 (5.7, 36.2) |
| *Missing* | *22* | *0.2%* | *0.6%* | 20 | 10.6 (6.6, 14.7) | 20 | 46.3 (17.5, 77.7) | 18 | 20.9 (17.8, 24.0) | 18 | 30.8 (5.6, 76.9) | 18 | 0.9 (0.0, 2.2) | 18 | 0.0 (0.0, 0.0) |
| **Sexual identity** |  |  |  |  |  |  |  |  |  |  |  |  |  |  |  |
| Heterosexual | 7872 | 81.4% | 81.2% | 7816 | 7.8 (7.6, 8.0) | 7817 | 17.4 (16.3, 18.7) | 7430 | 23.7 (23.5, 23.8) | 7427 | 15.8 (14.7, 17.1) | 7416 | 0.3 (0.3, 0.3) | 7445 | 10.0 (9.1, 11.1) |
| Gay or Lesbian | 409 | 4.2% | 4.2% | 408 | 10.2 (9.3, 11.1) | 408 | 33.0 (26.5, 40.1) | 390 | 22.5 (21.8, 23.2) | 385 | 31.2 (25.2, 37.8) | 384 | 0.7 (0.5, 0.8) | 392 | 17.9 (13.7, 23.1) |
| Bisexual | 914 | 9.4% | 9.2% | 914 | 12.0 (11.4, 12.5) | 914 | 43.2 (38.4, 48.2) | 884 | 21.3 (20.8, 21.7) | 886 | 46.3 (41.4, 51.3) | 880 | 1.0 (0.9, 1.1) | 891 | 33.7 (29.1, 38.7) |
| Other | 33 | 0.3% | 0.3% | 33 | 13.9 (12.3, 15.5) | 33 | 68.6 (48.6, 83.5) | 31 | 19.6 (18.4, 20.9) | 31 | 57.2 (36.5, 75.7) | 31 | 1.4 (0.8, 2.0) | 31 | 41.9 (23.1, 63.3) |
| Pansexual | 50 | 0.5% | 0.6% | 50 | 12.4 (10.7, 14.1) | 50 | 57.5 (31.8, 79.7) | 49 | 21.3 (19.6, 23.1) | 49 | 76.7 (57.6, 88.9) | 49 | 1.9 (0.9, 2.8) | 50 | 34.6 (17.2, 57.5) |
| Asexual | 48 | 0.5% | 0.4% | 48 | 11.4 (9.9, 13.0) | 48 | 37.0 (21.5, 55.8) | 48 | 21.7 (20.0, 23.4) | 47 | 50.4 (31.1, 69.5) | 46 | 1.1 (0.6, 1.7) | 46 | 16.3 (6.9, 34.0) |
| Queer | 25 | 0.3% | 0.2% | 25 | 10.7 (8.5, 12.9) | 25 | 38.4 (19.9, 60.9) | 25 | 22.5 (21.0, 24.0) | 25 | 41.6 (22.9, 63.1) | 25 | 1.1 (0.4, 1.8) | 25 | 19.0 (7.7, 39.9) |
| *Missing/Prefer not to say* | *324* | *3.3%* | *3.9%* | 55 | 11.0 (8.7, 13.2) | 55 | 39.8 (23.3, 59.0) | 48 | 19.6 (17.3, 21.9) | 50 | 29.9 (16.7, 47.5) | 50 | 0.6 (0.2, 0.9) | 63 | 24.5 (13.6, 40.1) |
| **History of childhood poverty** |  |  |  |  |  |  |  |  |  |  |  |  |  |  |  |
| No | 5889 | 60.9% | 64.4% | 5732 | 7.9 (7.8, 8.1) | 5733 | 17.8 (16.5, 19.2) | 5496 | 23.7 (23.6, 23.9) | 5487 | 19.9 (18.4, 21.4) | 5479 | 0.4 (0.3, 0.4) | 5505 | 10.7 (9.7, 11.8) |
| Yes | 3773 | 39.0% | 35.5% | 3605 | 9.2 (8.9, 9.5) | 3605 | 27.6 (25.3, 30.0) | 3397 | 22.5 (22.3, 22.8) | 3402 | 21.1 (19.0, 23.3) | 3391 | 0.5 (0.4, 0.5) | 3427 | 17.6 (15.6, 19.8) |
| *Missing* | *13* | *0.1%* | *0.1%* | 12 | 9.7 (4.4, 15.0) | 12 | 22.6 (5.5, 59.3) | 12 | 21.3 (16.1, 26.5) | 11 | 23.0 (5.6, 60.3) | 11 | 0.3 (0.0, 0.6) | 11 | 5.5 (0.7, 32.1) |

*Note.* * Weighted results; ** Results in cells with “-“ have been suppressed to preserve confidentiality. CI: confidence interval; N obs: number of observations; SWEMWBS: Short Warwick-Edinburgh Mental Well-Being Scale. Results for psychological distress (Kessler-6) and psychological wellbeing (SWEMWBS) are based on observed sum scores.

##### eFigure 9.1. Visualisation of weighted estimates by detailed ethnic identity/background levels (before recoding for analyses).

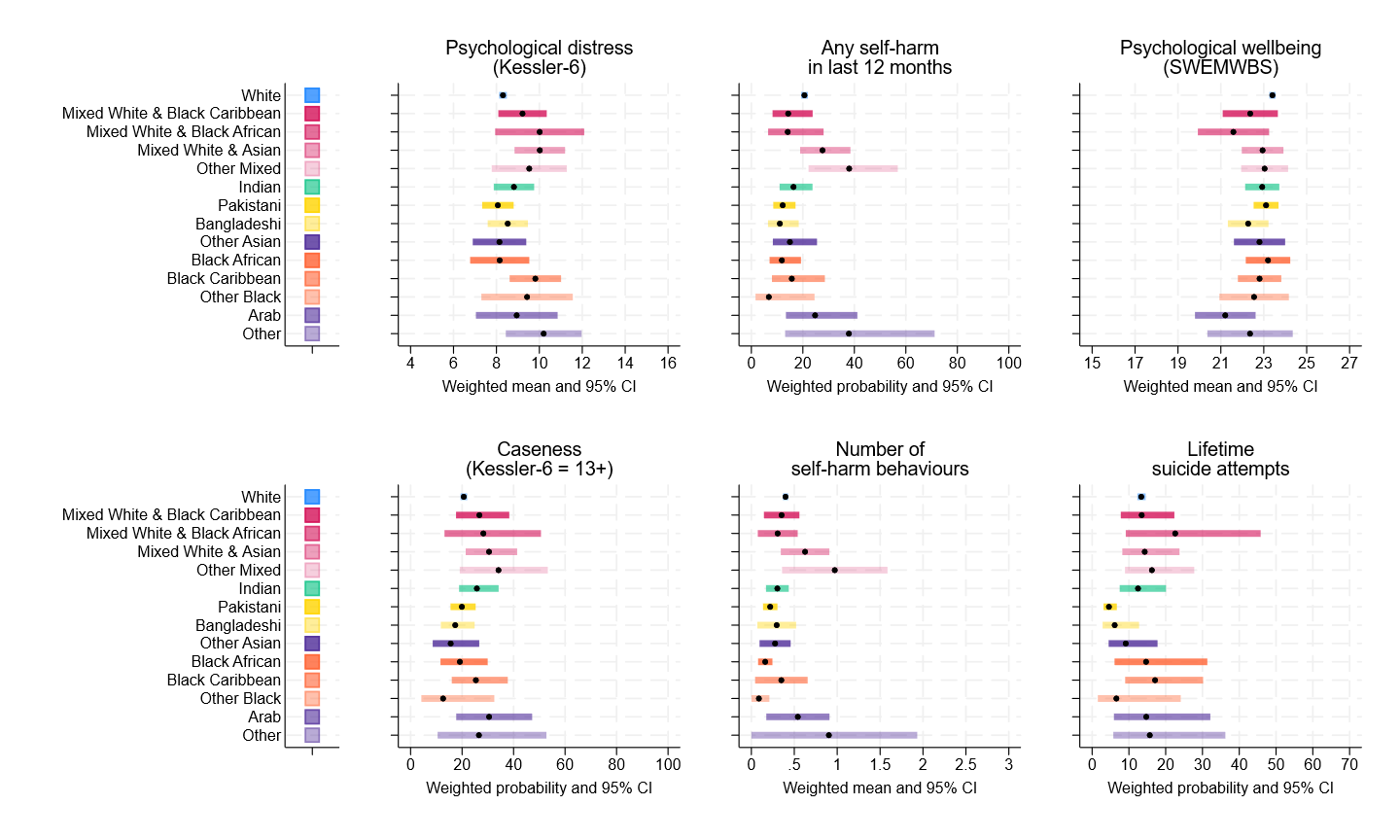

Note: results for Kessler-6 and Short Warwick Edinburgh Mental Well-Being Scale (SWEMWBS) are based on observed sum scores.

##### eFigure 9.2. Visualisation of weighted estimates by detailed sexual identity levels (before recoding for analyses.

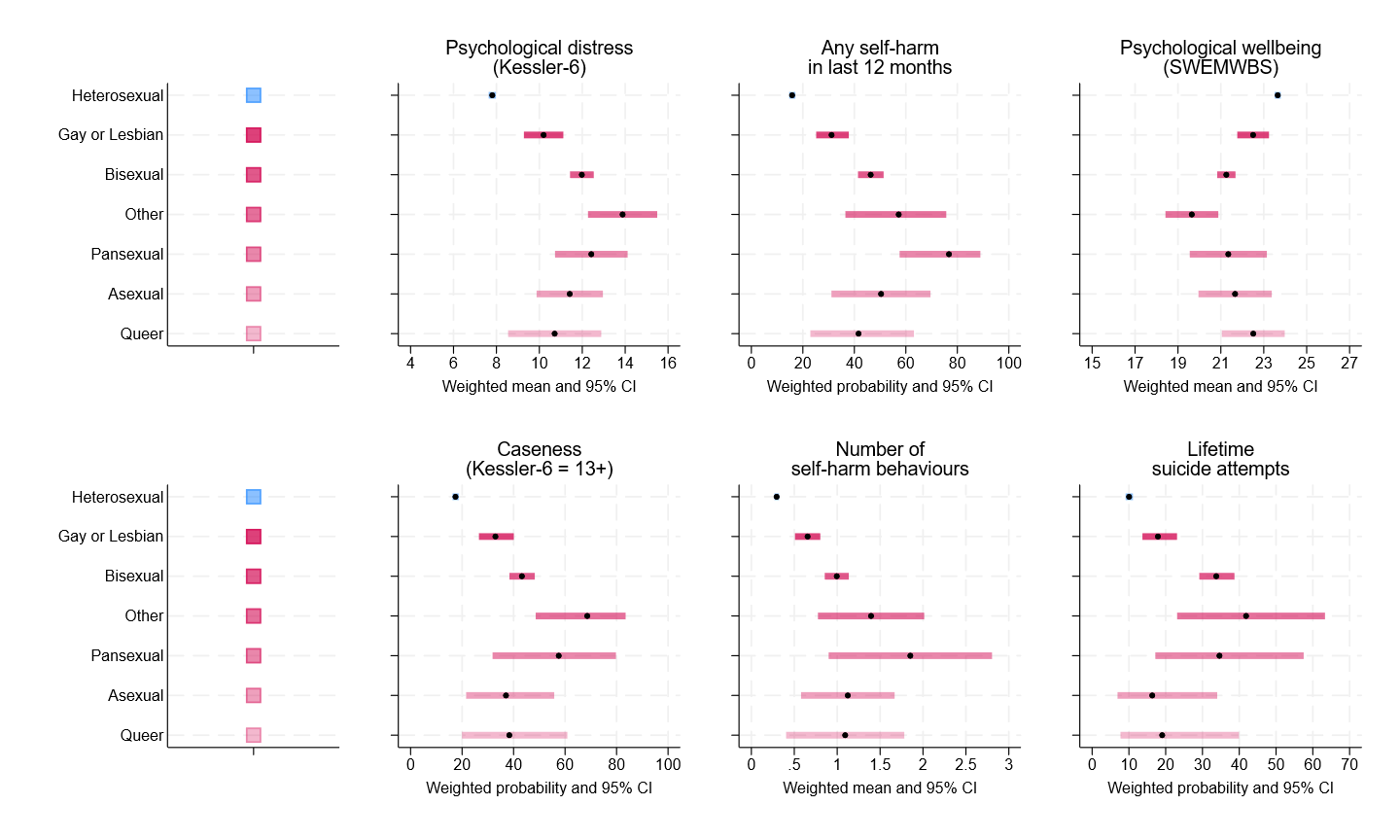

Note: results for Kessler-6 and Short Warwick Edinburgh Mental Well-Being Scale (SWEMWBS) are based on observed sum scores.

##### eFigure 9.3. Visualisation of weighted estimates across social identities and positions as used in the analyses.

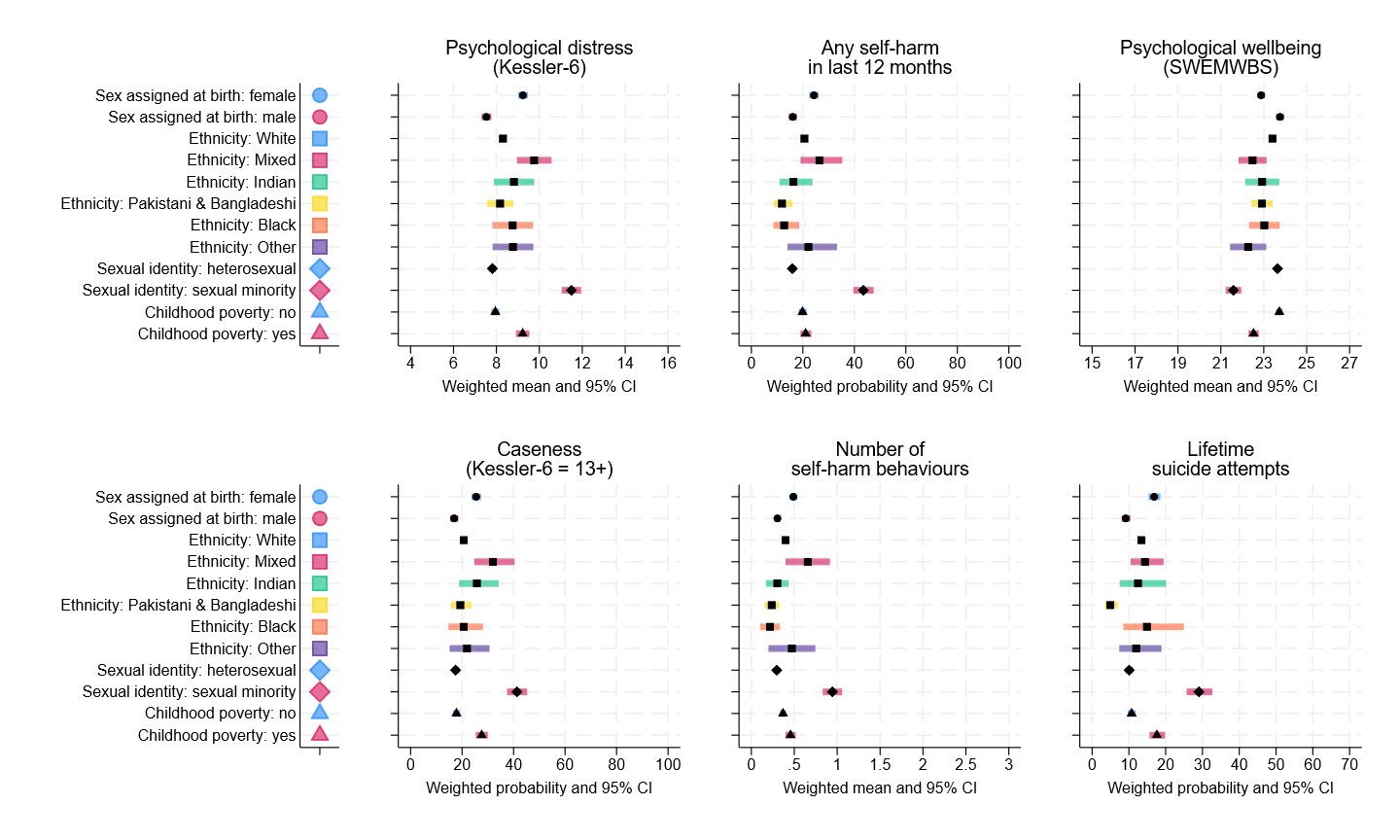

Note: results for Kessler-6 and Short Warwick Edinburgh Mental Well-Being Scale (SWEMWBS) are based on observed sum scores.

### 10. Detailed results from the measurement invariance testing

Configural models estimated in the entire sample showed acceptable yet not good/excellent model fit (**eTable 8.1**).

##### eTable 10.1. Results from configural models in entire sample

| **Measure** | **N** | **χ^2^ (df)** | **RMSEA (90% CI)** | **CFI** | **TLI** | **SRMR** |
| --- | --- | --- | --- | --- | --- | --- |
| Kessler-6 | 9390 | 561 (9) | 0.081 (0.075, 0.087) | 0.949 | 0.915 | 0.043 |
| SWEMWBS | 8939 | 429 (14) | 0.058 (0.053, 0.062) | 0.940 | 0.910 | 0.036 |

*Note.* χ^2^: chi-square; CFI: Comparative Fit Index; CI: confidence interval; df: degrees of freedom; N: number of observations; RMSEA: root mean squared error of approximation; SWEMWBS: Short Warwick-Edinburgh Mental Well-Being Scale; SRMR: Standardized Root Mean Square Residual; TLI: Tucker-Lewis Index.

#### 10.1. Results from the alignment method

An initial exploration with the alignment method suggested measurement invariance issues by sexual identity for the Kessler-6 (58.3% of non-invariant items) and by birth sex for the SWEMWBS (28.6% of non-invariant items) (**eTable 10.2**). The percentage of non-invariant items remained below the recommended 25% threshold in all other cases, including by ethnicity, childhood poverty, and by all the three-way combinations across the four social identities and positions (four-way combinations resulted in inappropriate solutions).

##### eTable 10.2. Results from the alignment method

|  | **Group / strata** | **N** | **Total number of groups / strata** | **Number of groups / strata excluded ^a^** | **Number of groups / strata included** | **Number of intercepts** | **N non-invariant intercepts** | **% non-invariant intercepts** | **N non-invariant loadings** | **% non-invariant loadings** | **Average % non-invariant items** |
| --- | --- | --- | --- | --- | --- | --- | --- | --- | --- | --- | --- |
| Kessler-6 | Birth sex | 9389 | 2 |  | 2 | 12 | 2 | 16.7% |  | 0.0% | 8.3% |
|  | Ethnicity | 9388 | 6 |  | 6 | 36 | 3 | 8.3% | 1 | 2.8% | 5.6% |
|  | **Sexual identity** | **9379** | **2** |  | **2** | **12** | **6** | **50.0%** | **8** | **66.7%** | **58.3%** |
|  | Childhood poverty | 9378 | 2 |  | 2 | 12 | 4 | 33.3% |  | 0.0% | 16.7% |
|  | Birth sex * ethnicity | 9388 | 12 |  | 12 | 72 | 9 | 12.5% | 1 | 1.4% | 6.9% |
|  | Birth sex * sexual identity | 9378 | 4 |  | 4 | 24 | 3 | 12.5% | 3 | 12.5% | 12.5% |
|  | Birth sex * childhood poverty | 9378 | 4 |  | 4 | 24 | 7 | 29.2% |  | 0.0% | 14.6% |
|  | Ethnicity * sexual identity | 9345 | 12 | 1 | 11 | 66 | 7 | 10.6% | 4 | 6.1% | 8.3% |
|  | Ethnicity * childhood poverty | 9253 | 12 | 1 | 11 | 66 | 5 | 7.6% |  | 0.0% | 3.8% |
|  | Sexual identity * childhood poverty | 9367 | 4 |  | 4 | 24 | 6 | 25.0% | 4 | 16.7% | 20.8% |
|  | Birth sex * sexual identity * childhood poverty | 9367 | 8 |  | 8 | 48 | 9 | 18.8% | 2 | 4.2% | 11.5% |
| SWEMWBS | **Birth sex** | **8938** | **2** |  | **2** | **14** | **8** | **57.1%** |  | **0.0%** | **28.6%** |
|  | Ethnicity | 8937 | 6 |  | 6 | 42 | 1 | 2.4% |  | 0.0% | 1.2% |
|  | Sexual identity | 8929 | 2 |  | 2 | 14 |  | 0.0% |  | 0.0% | 0.0% |
|  | Childhood poverty | 8927 | 2 |  | 2 | 14 |  | 0.0% |  | 0.0% | 0.0% |
|  | Birth sex * ethnicity | 8937 | 12 |  | 12 | 84 | 3 | 3.6% | 1 | 1.2% | 2.4% |
|  | Birth sex * sexual identity | 8928 | 4 |  | 4 | 28 | 4 | 14.3% |  | 0.0% | 7.1% |
|  | Birth sex * childhood poverty | 8927 | 4 |  | 4 | 28 | 4 | 14.3% |  | 0.0% | 7.1% |
|  | Ethnicity * sexual identity | 8878 | 12 | 3 | 9 | 63 | 0 | 0.0% |  | 0.0% | 0.0% |
|  | Ethnicity * childhood poverty | 8811 | 12 | 1 | 11 | 77 | 3 | 3.9% |  | 0.0% | 1.9% |
|  | Sexual identity * childhood poverty | 8917 | 4 |  | 4 | 28 | 1 | 3.6% |  | 0.0% | 1.8% |
|  | Birth sex * sexual identity * childhood poverty | 8917 | 8 |  | 8 | 56 | 6 | 10.7% |  | 0.0% | 5.4% |

*Note*. N: number of observations; SWEMWBS: Short Warwick-Edinburgh Mental Well-Being Scale. ^a^ Groups with less than 20 observations were excluded to prevent estimation issues and inappropriate solutions (e.g., negative variance estimates; for additional information, see pages 9-11 in this same Appendix). Rows highlighted in boldface correspond to the cases where alignment models suggested departures from for approximate measurement invariance, which were subsequently explored in additional detail using multiple-group confirmatory factor analysis (MG-CFA) models.

#### 10.2. Results from the multiple-group confirmatory factor analysis (MG-CFA) and partial measurement invariance models

Based on the results from the screening for approximate measurement invariance with the alignment models, multiple-group Confirmatory Factor Analysis (MG-CFA) measurement invariance models were estimated for the Kessler-6 by sexual identity and its intersections with birth sex, childhood poverty, and both; and for the SWEMWBS for birth sex and its intersections with sexual identity, childhood poverty, and both.

MG-CFA models initially showed no comparability issues across sexual identity groups for the Kessler-6: models with the level of measurement invariance required for mean comparisons (scalar invariance) across sexual identity groups showed acceptable/reasonable (though not good/excellent) fit. However, models at the intersection of sexual identity and birth sex, childhood poverty, and both consistently showed issues in the measurement invariance, requiring freeing certain measurement parameters.

At the most granular level tested (birth sex * sexual identity * childhood poverty), measurement comparability issues with the Kessler-6 exclusively concerned items 6 (“nervous”), 3 (“restless/fidgety”), and 4 (“everything was an effort”). On average and at the same underlying distress levels, heterosexual males with and without a childhood poverty history provided lower-than-expected responses for item 6 (“nervous”), whilst females without a childhood poverty history and regardless of their sexual identity provided higher-than-expected responses; females without a childhood poverty history provided lower-than-expected responses for item 3 (“restless/fidgety”) if from a sexual minority, and higher-than-expected responses if heterosexual; and item 4 (“everything was an effort”) was a comparatively poorer indicator of psychological distress for sexual minority females, who, on average, also provided higher-than-expected responses at the same underlying distress levels.

At the most granular level tested (birth sex * sexual identity * childhood poverty), measurement comparability issues with the SWEMWBS exclusively concerned heterosexual females with no childhood poverty history. On average and at the same underlying wellbeing levels, they provided higher-than-expected responses to items 1 (“feeling optimistic about the future”) and 6 (“feeling close to other people”), and lower-than-expected responses to item 7 (“able to make up my own mind”).

**eTable 10.3** includes the detailed results from the model comparison approach implemented. Partially invariant Kessler-6 and SWEMWBS factor scores were derived from the most granular and invariant models to account for the identified measurement differences across groups.

##### eTable 10.3. Results from multiple-group confirmatory factor analysis (MG-CFA) models

|  | **Group / strata** | **N** | **Model** | **χ^2^ (df)** | **RMSEA** | **CFI** | **TLI** | **Diff  RMSEA** | **Diff  CFI** | **Free parameters** |
| --- | --- | --- | --- | --- | --- | --- | --- | --- | --- | --- |
| Kessler-6 | Sexual identity | 9379 | Configural | 599 (18) | 0.083 | 0.950 | 0.917 |  |  |  |
|  |  |  | Metric | 692 (23) | 0.079 | 0.943 | 0.925 | -0.004 | 0.007 |  |
|  |  |  | Scalar | 744 (28) | 0.074 | 0.939 | 0.934 | -0.005 | 0.004 |  |
|  | Birth sex *  sexual identity | 9378 | Configural | 627 (36) | 0.084 | 0.950 | 0.916 |  |  |  |
|  |  |  | Metric | 719 (51) | 0.075 | 0.943 | 0.933 | -0.009 | 0.007 |  |
|  |  |  | Scalar | 936 (66) | 0.075 | 0.926 | 0.933 | 0.000 | 0.017 |  |
|  |  |  | Scalar (partial) | 794 (64) | 0.070 | 0.938 | 0.942 | -0.005 | 0.005 | Intercept 6 in strata 10 (female + heterosexual) and 20 (male + heterosexual) |
|  | Sexual identity *  childhood poverty | 9367 | Configural | 587 (36) | 0.081 | 0.949 | 0.915 |  |  |  |
|  |  |  | Metric | 719 (51) | 0.075 | 0.938 | 0.927 | -0.006 | 0.011 |  |
|  |  |  | Metric (partial) | 690 (50) | 0.074 | 0.941 | 0.929 | -0.007 | 0.008 | Loading 4 in stratum 11 (sexual minority + childhood poverty) |
|  |  |  | Metric (partial) Scalar | 843 (65) | 0.071 | 0.928 | 0.934 | -0.003 | 0.013 | Loading 4 in stratum 11 (sexual minority + childhood poverty) |
|  |  |  | Metric (partial) Scalar (partial) | 798 (64) | 0.070 | 0.932 | 0.937 | -0.004 | 0.009 | Loading 4 in stratum 11 (sexual minority + childhood poverty) Intercept 6 in stratum 01 (heterosexual + childhood poverty) |
|  | **Birth sex *  sexual identity *  childhood poverty** | 9367 | Configural | 665 (72) | 0.084 | 0.949 | 0.916 |  |  |  |
|  |  |  | Metric | 825 (107) | 0.076 | 0.939 | 0.931 | -0.008 | 0.010 |  |
|  |  |  | Metric (partial) | 798 (106) | 0.075 | 0.941 | 0.933 | -0.009 | 0.008 | Loading 4 in stratum 111 (female + sexual minority + childhood poverty) |
|  |  |  | Metric (partial) Scalar | 1140 (141) | 0.078 | 0.915 | 0.927 | 0.003 | 0.026 | Loading 4 in stratum 111 (female + sexual minority + childhood poverty) |
|  |  |  | **Metric (partial) Scalar (partial)** | **899 (134)** | **0.070** | **0.935** | **0.942** | **-0.005** | **0.006** | Loading 4 [**0.866**] in stratum 111 [0.553] (female + sexual minority + childhood poverty) Intercept 6 [**1.657**] in strata 100 [1.843] (female + heterosexual + no childhood poverty), 110 [1.895] (female + sexual minority + no childhood poverty), 200 [1.513] (male + heterosexual + no childhood poverty), and 201 [1.322] (male + heterosexual + childhood poverty) Intercept 3 [**1.747**] in strata 101 [1.612], 110 [1.974] (female + heterosexual + childhood poverty)  Intercept 4 [**1.601**] in stratum 111 [2.033] (female + sexual minority + childhood poverty) |
| SWEMWBS | Birth sex | 8938 | Configural | 456 (28) | 0.058 | 0.937 | 0.906 |  |  |  |
|  |  |  | Metric | 479 (34) | 0.054 | 0.935 | 0.920 | -0.004 | 0.002 |  |
|  |  |  | Scalar | 586 (40) | 0.055 | 0.920 | 0.916 | 0.001 | 0.015 |  |
|  |  |  | Scalar (partial) | 528 (39) | 0.053 | 0.929 | 0.923 | -0.001 | 0.006 | Intercept 6 across both groups |
|  | Birth sex *  sexual identity | 8928 | Configural | 499 (56) | 0.060 | 0.937 | 0.906 |  |  |  |
|  |  |  | Metric | 539 (74) | 0.053 | 0.934 | 0.925 | -0.007 | 0.003 |  |
|  |  |  | Scalar | 666 (92) | 0.053 | 0.919 | 0.926 | 0.000 | 0.015 |  |
|  |  |  | Scalar (partial) | 610 (91) | 0.051 | 0.927 | 0.932 | -0.002 | 0.007 | Intercept 6 in stratum 20 (male + heterosexual) |
|  | Birth sex *  childhood poverty | 8927 | Configural | 468 (56) | 0.057 | 0.937 | 0.906 |  |  |  |
|  |  |  | Metric | 520 (74) | 0.052 | 0.932 | 0.923 | -0.005 | 0.005 |  |
|  |  |  | Scalar | 682 (92) | 0.054 | 0.910 | 0.918 | 0.002 | 0.022 |  |
|  |  |  | Scalar (partial) | 589 (90) | 0.050 | 0.924 | 0.929 | -0.002 | 0.008 | Intercept 6 and 7 in stratum 10 (female + no childhood poverty) |
|  | **Birth sex *  sexual identity *  childhood poverty** | 8917 | Configural | 559 (112) | 0.060 | 0.937 | 0.906 |  |  |  |
|  |  |  | Metric | 645 (154) | 0.054 | 0.931 | 0.925 | -0.006 | 0.006 |  |
|  |  |  | Scalar | 858 (196) | 0.055 | 0.907 | 0.920 | 0.001 | 0.024 |  |
|  |  |  | **Scalar (partial)** | **738 (193)** | **0.050** | **0.923** | **0.933** | **-0.004** | **0.008** | Intercepts 1 [3.486 vs **3.320**], 6 [3.607 vs **3.402**] and 7 [3.701 vs **3.849**] in stratum 100 (female + heterosexual + no childhood poverty) |

*Note.* χ^2^: chi-square; CFI: Comparative Fit Index; df: degrees of freedom; Diff: difference; N: number of observations; RMSEA: root mean squared error of approximation; SWEMWBS: Short Warwick-Edinburgh Mental Well-Being Scale; TLI: Tucker-Lewis Index. Models highlighted in boldface correspond to the most invariant and granular models achieved for each of the measures; additional details on the specific values of the non-invariant measurement parameters are provided for those models, with the values of the parameters for most groups in boldface and underlined, and the values of the freed parameters in the specific groups in blue or red for parameter values significantly higher or lower than the invariant ones.

### 12. Visual comparison of stratum-specific estimates across sensitivity analyses.

The following pages include the visualisation of stratum-specific estimates across the different sensitivity analyses: models with observed sum scores (**eFigure 11.1**), models excluding participants from the mop-up survey (**eFigure 11.2**), models with the number of self-harm behaviours in the past 12 months (**eFigure 11.3**), and quantitative bias analysis (QBA) models on mode and selection effects (**eFigures 11.4-11.7**).

Across all plots, dots represent predicted outcome levels (means for Kessler-6 and SWEMWBS, prevalence estimates for ‘caseness’, self-harm, and lifetime suicide attempts) or random (non-additive) effects (expressed as the difference in mean or prevalence estimates compared to the expectation based on additive effects alone), and accompanying lines represent the 95% confidence intervals. Random effects significantly different than zero (not overlapping with zero) are highlighted in red.

Intersectional strata are coded using four digits, each representing one variable. First digit: sex assigned at birth (1 female, 2 male); second digit: ethnic identity/background (1 White, 2 Mixed, 3 Indian, 4 Pakistani/Bangladeshi, 5 Black, 6 Other); third digit: sexual identity (0 Heterosexual, 1 Sexual minority); fourth digit: history of childhood poverty (0 No, 1 Yes).

All results are weighted using combined survey and non-response weights, and results from strata with less than 10 observations are not shown to preserve confidentiality.

##### eFigure 11.1. Caterpillar plots of predicted outcome levels and random (non-additive) effects across intersectional strata using observed Kessler-6 and Short Warwick Edinburgh Well-Being Scale (SWEMWBS) sum scores; weighted results.

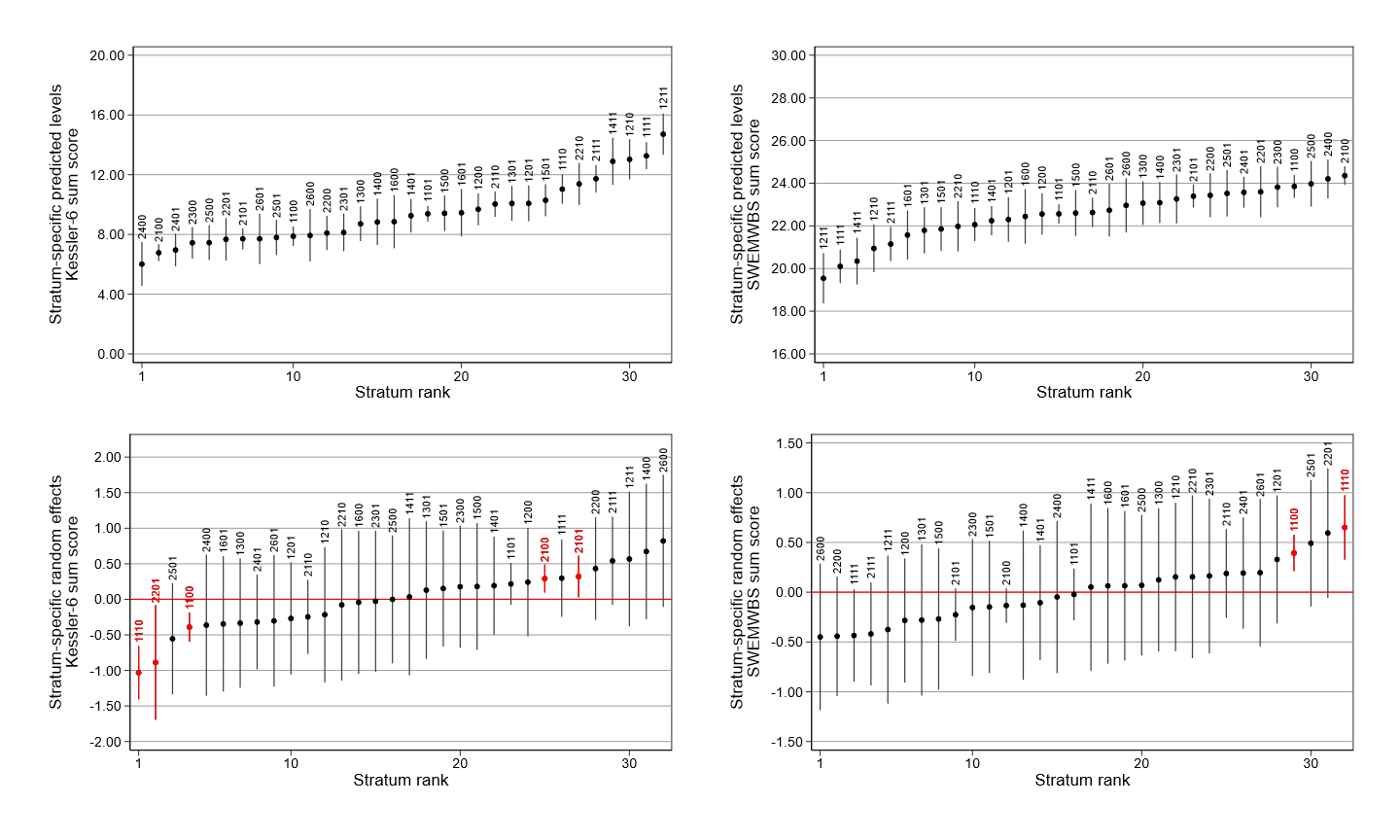

##### eFigure 11.2. Caterpillar plots of predicted outcome levels and random (non-additive) effects across intersectional strata excluding participants from mop-up survey; weighted results.

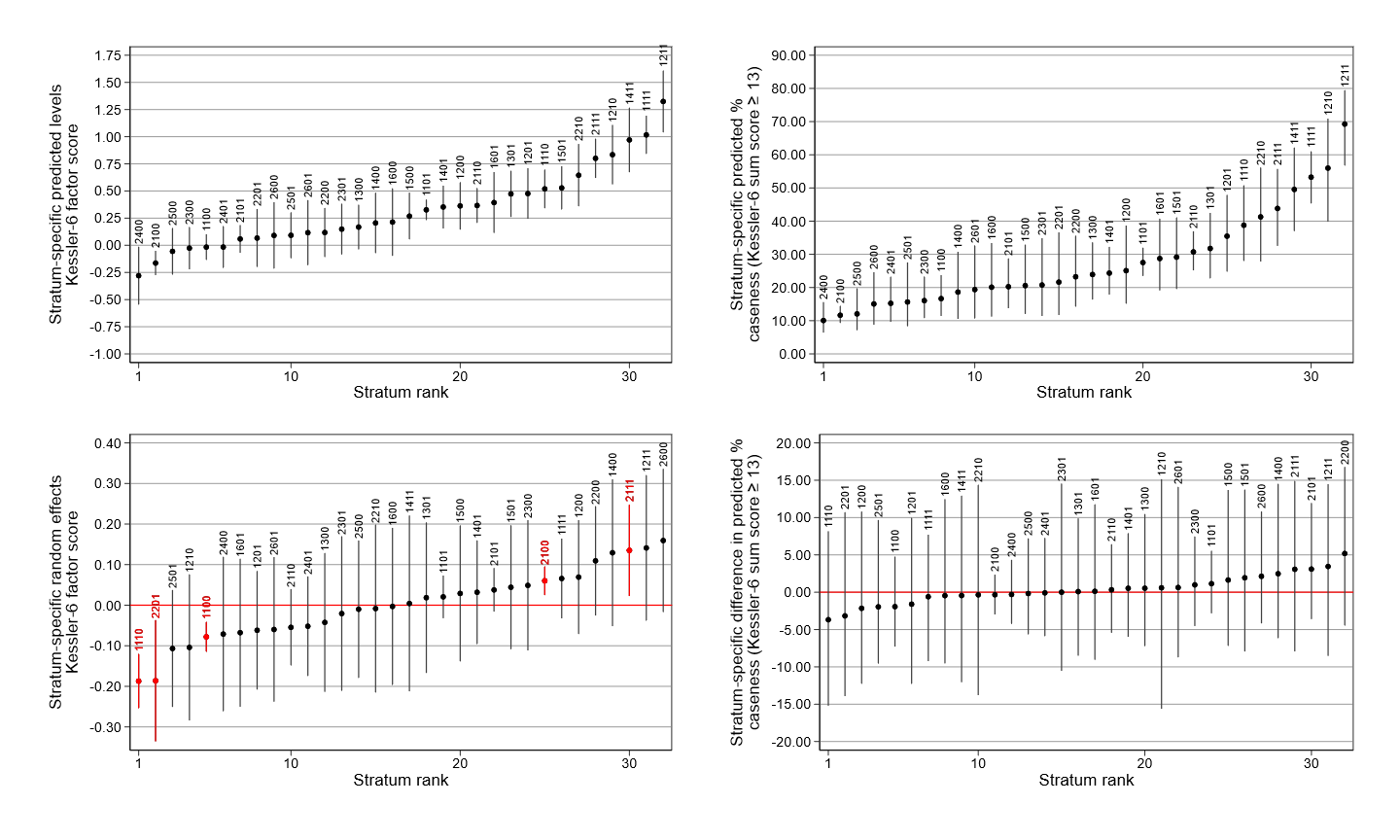

##### eFigure 11.3. Caterpillar plots of predicted outcome levels and random (non-additive) effects across intersectional strata using number of self-harm behaviours in the past 12 months; weighted results.

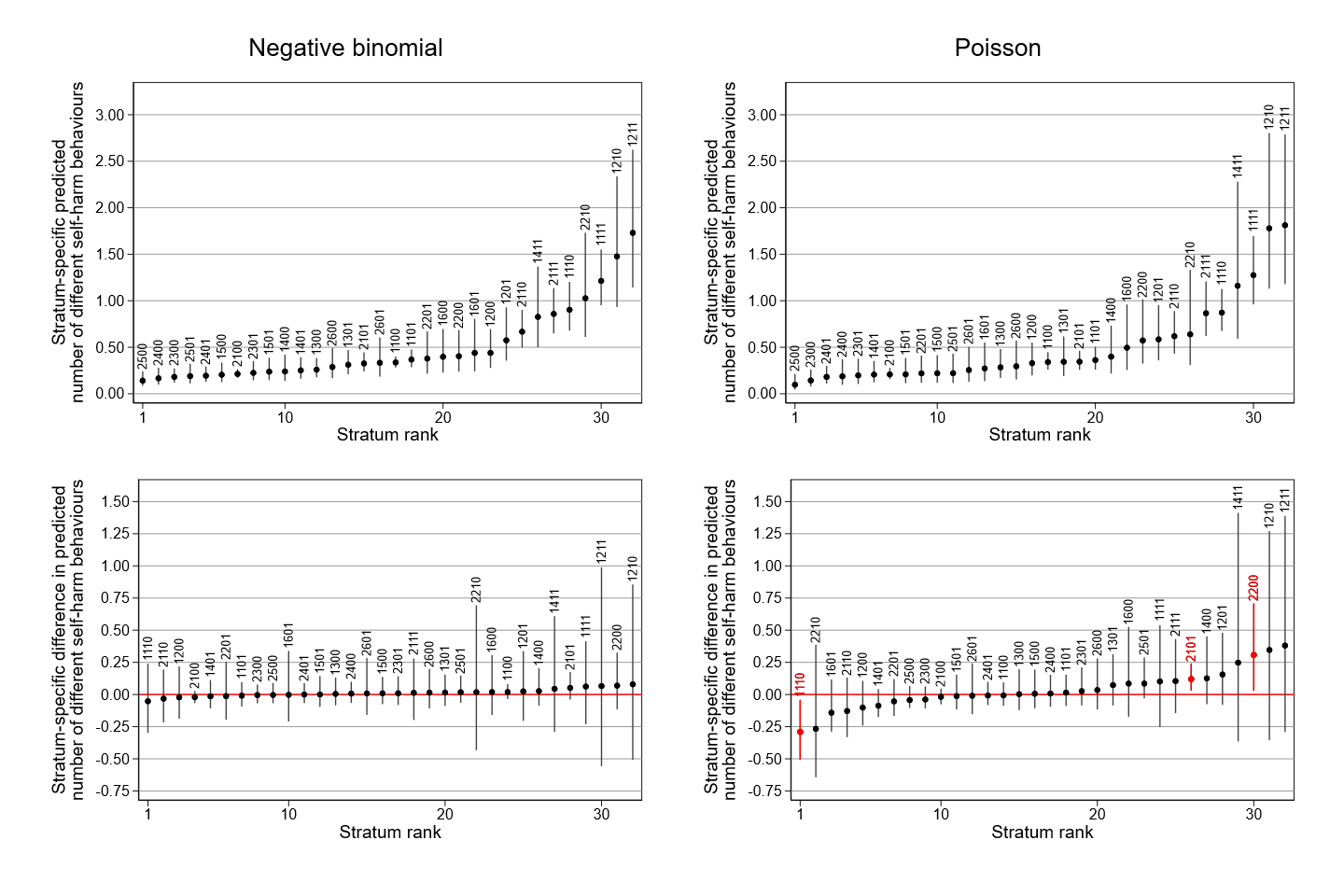

##### eFigure 11.4. Caterpillar plots of predicted outcome levels across intersectional strata across quantitative bias analysis (QBA) scenarios, Kessler-6 factor scores; weighted results.

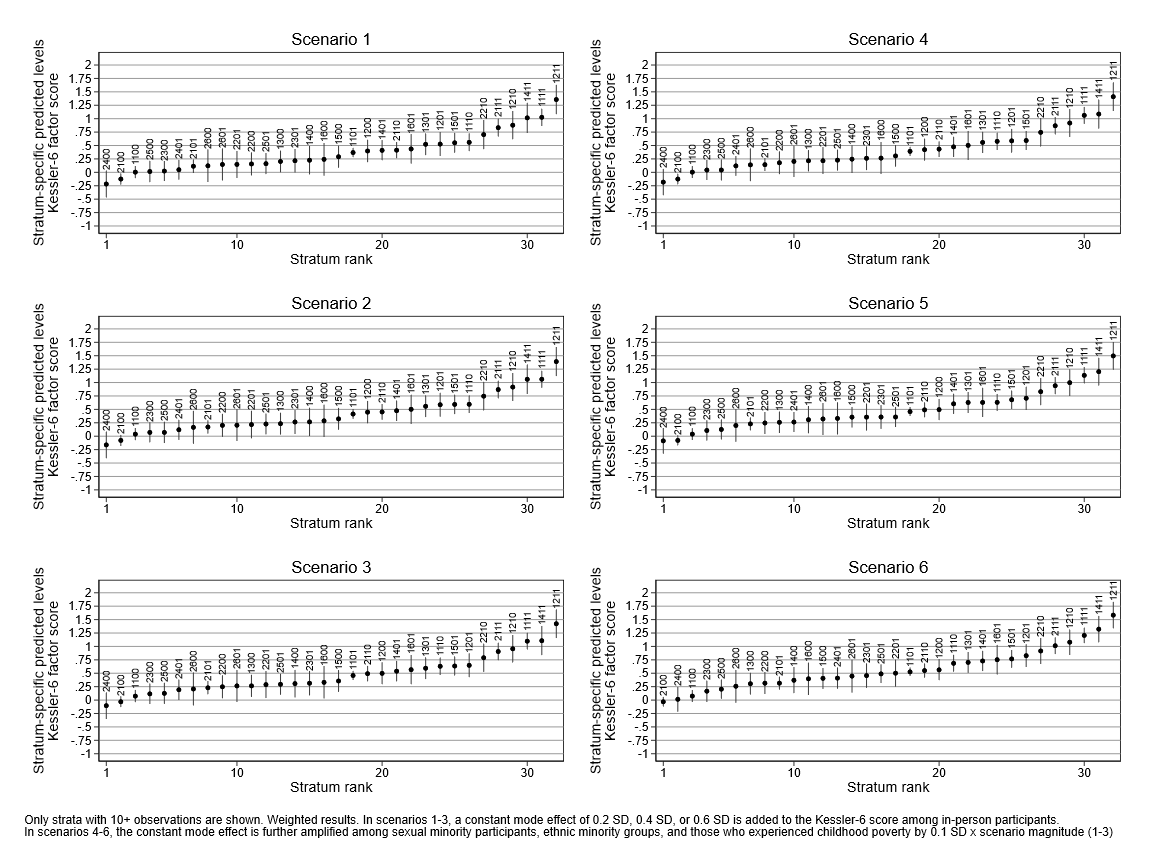

##### eFigure 11.5. Caterpillar plots of random (non-additive) effects across intersectional strata across quantitative bias analysis (QBA) scenarios, Kessler-6 factor scores; weighted results.

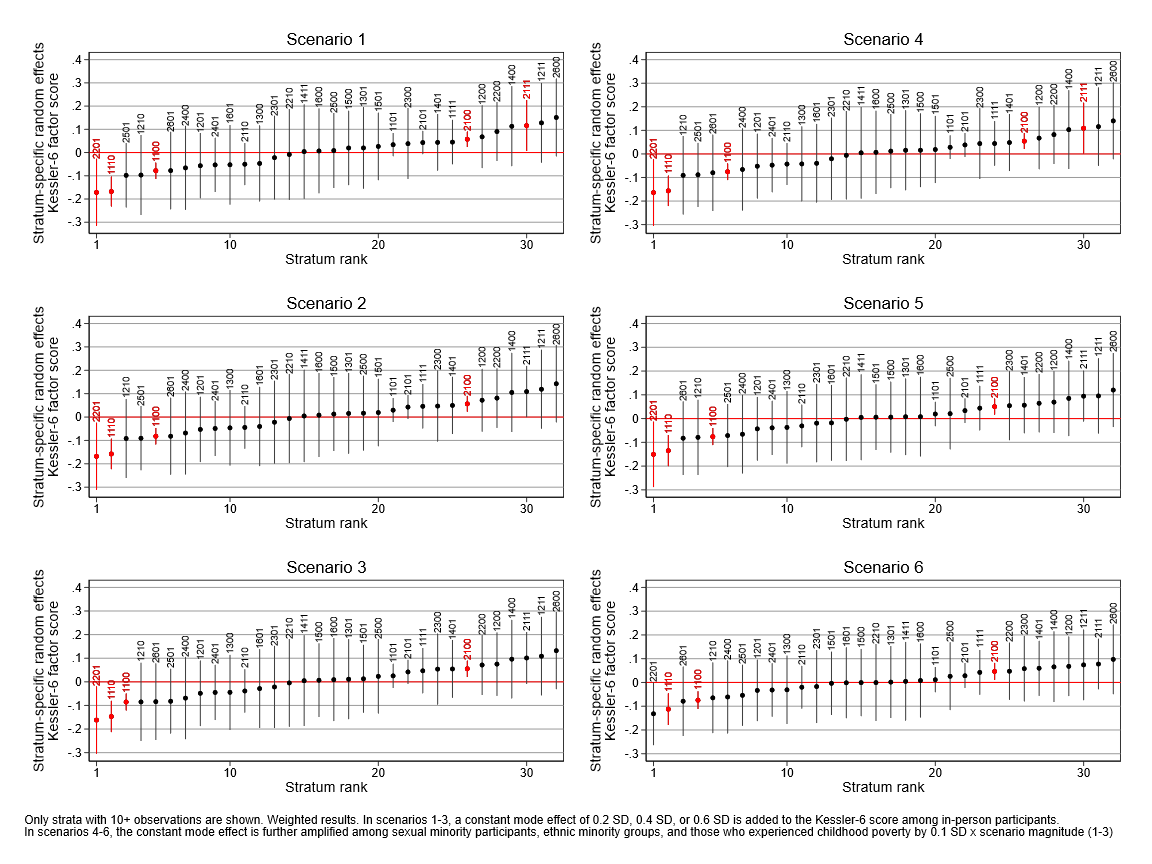

##### eFigure 11.6. Caterpillar plots of predicted outcome levels across intersectional strata across quantitative bias analysis (QBA) scenarios, SWEMWBS factor scores; weighted results.

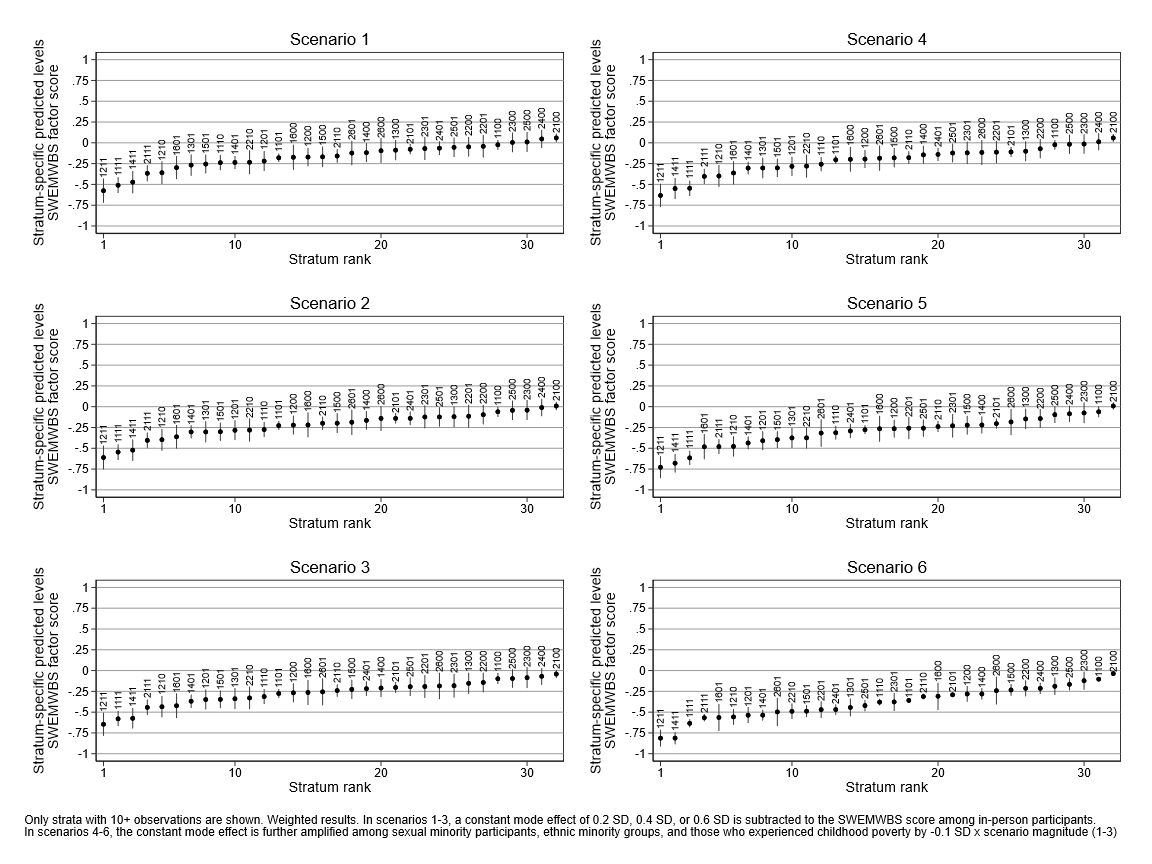

##### eFigure 11.7. Caterpillar plots of random (non-additive) effects across intersectional strata across quantitative bias analysis (QBA) scenarios, SWEMWBS factor scores; weighted results.

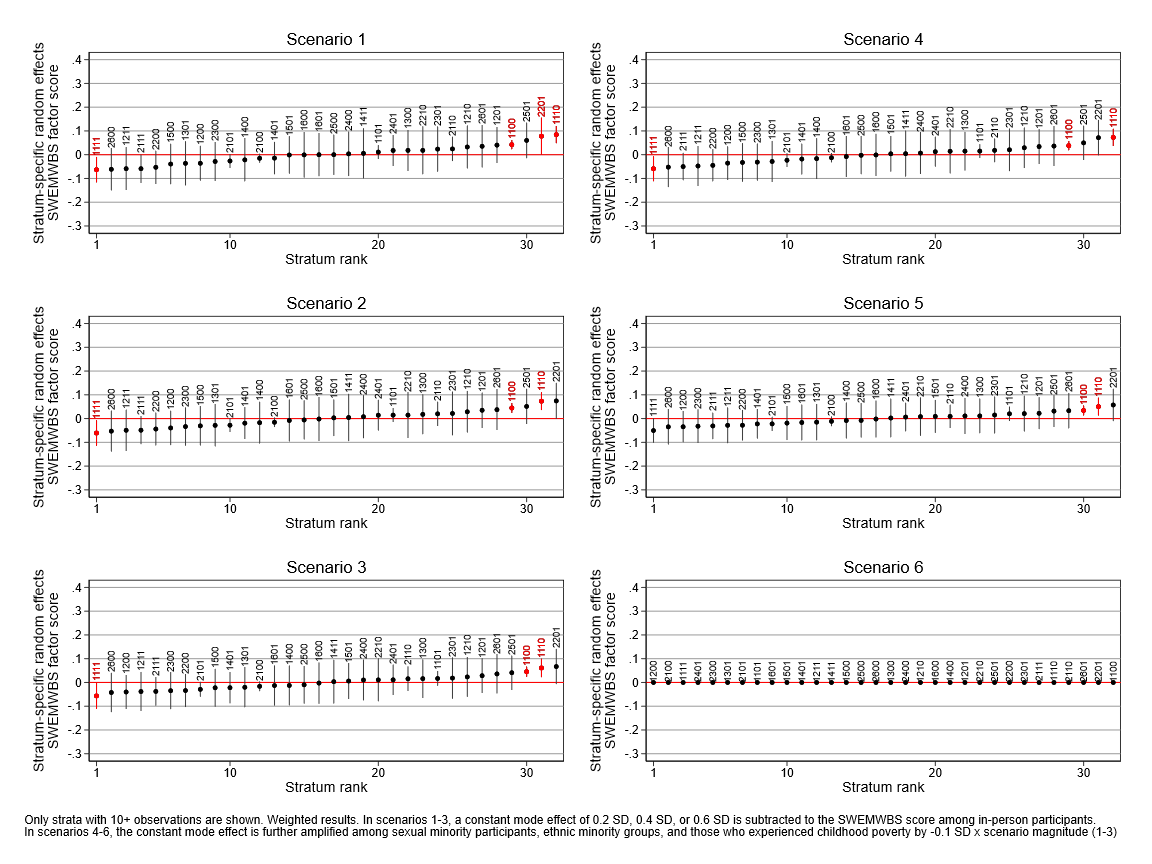

### 13. Silences and absences: who is not (sufficiently) represented in this study

Some young people within the UK population are not included in this study. Not by accident, in many of the cases they are among the most marginalised young members of society. We would like to make them visible even through the explicit acknowledgement of their absence.

In some cases, they are not included because they were not part of the issued sample of the Millennium Cohort Study. To be eligible, infants had to be included in the Child Benefit register, a universal benefit at the time that virtually captured all of the child population “except those ineligible due to recent or temporary immigrant status”.^27^ This would likely include children of asylum seekers, refugees without settled status, or other recent migrants who did not appear on the sampling list. The sampling also excluded infants who had already been placed in foster/institutional care by the age of 9 months. In other words, infants who were not living with a private family unit at 9 months (e.g., those in institutional care or adoptive placement by that age) would have not been included. Additionally, the requirement of being resident in the sampled area at 9 months likely means that some highly mobile or transient families (including, for instance, those experiencing homelessness or those from Traveller communities) may have also been missed, since they may not have appeared on local records or have stable addresses in the selected areas at the required time.^28^ We were also unable to include in our study individuals that may not have been able to take part on the age 23 sweep (for instance, young people who were imprisoned or without a home and the means to be contacted by the survey team at the time of data collection, including victims of modern slavery). Although all these groups likely concern a very small fraction of the population, they do also involve some of the most marginalised groups, who are thus invisible in this study’s data. The use of a cohort of people born in the UK also inherently means that first generation migrants are not represented in our results.

In other cases, they are not included or they are grouped together with other categories (as is the case with multiple ethnic and sexual identity groups) because, due to the available data and the aims of our study (to understand how multiple systems of oppression intersect and produce unequal mental health outcomes in young people in the UK), we were not able to meaningfully engage with the research question at hand with a larger degree of granularity. Pooling together some of these categories may be masking underlying differences and inequalities. In the case of sexual identity, and as also shown in the granular descriptive analysis conducted before recoding, sexual minority identities show a gradient of worse results compared to heterosexual individuals. However, the intersection of the more granular sexual identity categories with other identities and positions may have shown different relationships. With regards to ethnic identity/background, some evidence shows that some groups (for instance, individuals with different Mixed backgrounds)^29^ may experience different degrees of discrimination, which in turn may differentially impact their mental health outcomes. Limited by the sample size, we aimed to retain as much granularity as possible, also respecting the priorities by the YPAG shaping this study and broader project, and acknowledged those underlying differences by providing population-wide estimates at each of those single yet more granular ethnic and sexual identities (see pages 17-22 in this same Appendix). However, by combining together these different groups, we may have flattened that complexity.

Our study also focused on a selected and definitely not exhaustive number of social identities and positions representing an also non-exhaustive number of systems of oppression. The methods we used and the available data meant we were not able to represent other intersecting systems of oppression whose impacts can be crucial for youth mental health, including, for example, transphobia, ableism, and religious discrimination.

### 14. Additional references

1.	Kessler RC, Green JG, Gruber MJ, et al. Screening for serious mental illness in the general population with the K6 screening scale: results from the WHO World Mental Health (WMH) survey initiative. *International journal of methods in psychiatric research* 2010; **19 Suppl 1**(Suppl 1): 4–22.

2.	Prochaska JJ, Sung HY, Max W, Shi Y, Ong M. Validity study of the K6 scale as a measure of moderate mental distress based on mental health treatment need and utilization. *International journal of methods in psychiatric research* 2012; **21**(2): 88–97.

3.	Kessler RC, Barker PR, Colpe LJ, et al. Screening for serious mental illness in the general population. *Archives of general psychiatry* 2003; **60**(2): 184–9.

4.	Stewart-Brown S, Tennant A, Tennant R, Platt S, Parkinson J, Weich S. Internal construct validity of the Warwick-Edinburgh Mental Well-being Scale (WEMWBS): a Rasch analysis using data from the Scottish Health Education Population Survey. *Health Qual Life Outcomes* 2009; **7**: 15.

5.	McAra L, McVie S. Youth crime and justice: Key messages from the Edinburgh Study of Youth Transitions and Crime. *Criminol Crim Justic* 2010; **10**(2): 179–209.

6.	McCall L. The complexity of intersectionality. *Signs* 2005; **30**(3): 1771–800.

7.	Brown M, Fitzsimons E, Haselden L, et al. Millennium Cohort Study Sweep 8: Age 23 Survey User Guide (Version 2). London: UCL Centre for Longitudinal Studies; 2026.

8.	Tabor E, Kneale D, Patalay P. Mainly heterosexual, bisexual, or other?: The measurement of sexual minority status and its impact on analytic sample, demographic distribution and health outcomes. *PloS one* 2024; **19**(9): e0303100.

9.	Chua YW, Schluter D, Pearce A, Sharp H, Taylor-Robinson D. Socioeconomic inequalities in mental health difficulties over childhood: a longitudinal sex-stratified analysis using the UK Millennium Cohort Study. *Soc Sci Med* 2025; **378**: 118159.

10.	Straatmann VS, Lai E, Lange T, et al. How do early-life factors explain social inequalities in adolescent mental health? Findings from the UK Millennium Cohort Study. *J Epidemiol Community Health* 2019; **73**(11): 1049–60.

11.	Rohrer JM, Paulewicz B. Rethinking measurement invariance causally. *Curr Res Ecol Soc Ps* 2025; **9**.

12.	Else-Quest NM, Hyde JS. Intersectionality in Quantitative Psychological Research: II. Methods and Techniques. *Psychol Women Quart* 2016; **40**(3): 319–36.

13.	Gregorich SE. Do self-report instruments allow meaningful comparisons across diverse population groups? Testing measurement invariance using the confirmatory factor analysis framework. *Med Care* 2006; **44**(11 Suppl 3): S78–94.

14.	Asparouhov T, Muthén B. Multiple-Group Factor Analysis Alignment. *Structural Equation Modeling-a Multidisciplinary Journal* 2014; **21**(4): 495–508.

15.	Luong R, Flake JK. Measurement invariance testing using confirmatory factor analysis and alignment optimization: A tutorial for transparent analysis planning and reporting. *Psychol Methods* 2023; **28**(4): 905–24.

16.	Hu LT, Bentler PM. Cutoff Criteria for Fit Indexes in Covariance Structure Analysis: Conventional Criteria Versus New Alternatives. *Structural Equation Modeling-a Multidisciplinary Journal* 1999; **6**(1): 1–55.

17.	Muthen B, Asparouhov T. IRT studies of many groups: the alignment method. *Front Psychol* 2014; **5**: 978.

18.	Chen FF. Sensitivity of goodness of fit indexes to lack of measurement invariance. *Structural Equation Modeling-a Multidisciplinary Journal* 2007; **14**(3): 464–504.

19.	Evans CR, Williams DR, Onnela JP, Subramanian SV. A multilevel approach to modeling health inequalities at the intersection of multiple social identities. *Soc Sci Med* 2018; **203**: 64–73.

20.	Evans CR, Leckie G, Subramanian SV, Bell A, Merlo J. A tutorial for conducting intersectional multilevel analysis of individual heterogeneity and discriminatory accuracy (MAIHDA). *SSM Popul Health* 2024; **26**: 101664.

21.	Leckie G, Browne WJ, Goldstein H, Merlo J, Austin PC. Partitioning variation in multilevel models for count data. *Psychol Methods* 2020; **25**(6): 787–801.

22.	Hancock A-M. Empirical intersectionality: A tale of two approaches. *UC Irvine Law Review* 2013; **3**: 259–96.

23.	Van Dusen B, Cian H, Nissen J, Arellano L, Woods AD. Comparing the Efficacy of Fixed-Effects and MAIHDA Models in Predicting Outcomes for Intersectional Social Strata. *Sociol Educ* 2024; **97**(4): 342–62.

24.	Goodman A, Brown M, Silverwood RJ, et al. The impact of using the Web in a mixed-mode follow-up of a longitudinal birth cohort study: Evidence from the National Child Development Study. *J Roy Stat Soc A* 2022; **185**(3): 822–50.

25.	Rickwood DJ, Coleman-Rose CL. The effect of survey administration mode on youth mental health measures: Social desirability bias and sensitive questions. *Heliyon* 2023; **9**(9): e20131.

26.	Tomova GD, Silverwood RJ, Tennant PWG, Wright L. How can the use of different modes of survey data collection introduce bias? An introduction to mode effects using directed acyclic graphs (DAGs). *Am J Epidemiol* 2026; **195**(5): 1406–15.

27.	Centre for Longitudinal Studies. Millennium Cohort Study User Guide (Surveys 1-5) (9th Edition). London: UCL Centre for Longitudinal Studies; 2020.

28.	Fitzsimons E, Pople L, Libuy N, Calderwood L, Haselden L. Reflections on the Design and Development of First 18 Years of the Millennium Cohort Study. *Eur Psychol* 2025; **30**(3): 148–60.

29.	Lam J, Koay A, Cortina-Borja M, Aldridge R, Blackburn R, Harron K. Intersectional relationships between age, sex, ethnicity, nationality and experience of racism in the UK using different ethnicity categorisations: A comparative study using survey data. *J Migr Health* 2026; **13**: 100384.
